# Spatiotemporal Tau Accumulation and Its Clinical Impact on PSP: Longitudinal Florzolotau (18F) PET

**DOI:** 10.64898/2026.04.26.26350467

**Authors:** Yoshikazu Chishiki, Kenji Tagai, Yuko Kataoka, Ryoji Goto, Kenta Osawa, Asaka Oyama, Hideki Matsumoto, Masanori Ichihashi, Yuki Momota, Tetsuji Kamada, Chie Seki, Kiwamu Matsuoka, Kosei Hirata, Shin Kurose, Sho Moriguchi, Yuki Komatsu, Hideo Kato, Yasuharu Yamamoto, Yoshikazu Nakano, Shigeki Hirano, Hitoshi Shinotoh, Hitoshi Shimada, Tokahiko Tokuda, Kazunori Kawamura, Ming-Rong Zhang, Keisuke Takahata, Makoto Higuchi, Hironobu Endo

## Abstract

**Background:** Florzolotau (18F) positron emission tomography (florzolotau PET) enables high-contrast *in vivo* detection of four-repeat tau pathology in progressive supranuclear palsy (PSP), but whether longitudinal tau imaging reflects disease progression remains unclear.

**Objectives:** To explore longitudinal tau accumulation using florzolotau PET and evaluate its association with clinical progression in PSP.

**Methods:** Twenty-six patients with PSP (18 Richardson’s syndrome [PSP-RS], 8 non-RS) and 10 age- and sex-matched healthy controls (HCs) underwent florzolotau PET, MRI, and clinical assessments at baseline and after 1 year. Regional standardized uptake value ratios (SUVR) were extracted across 57 regions of interest. Partial least squares (PLS) multivariate analyses revealed regions with elevated baseline SUVR and longitudinal increase (ΔSUVR) in PSP relative to HCs, alongside regions where ΔSUVR was associated with changes in PSP Rating Scale scores.

**Results:** At baseline, tau deposition was most prominent in the globus pallidus (GP) and midbrain in patients with PSP. Longitudinal tau increases were observed in the GP, frontoparietal cortex, and cerebellar white matter, and minimal progression was observed in the midbrain. GP tau accumulation exhibited the strongest association with clinical progression in the PLS model among PSP-RS and a univariate correlation (Spearman’s ρ = 0.688, *p* = 0.002).

**Conclusions:** This study provides *in vivo* evidence of the spatiotemporal progression of tau pathology in PSP. In the GP, tau accumulation emerges early and continues to increase with clinical deterioration. These findings support the utility of florzolotau PET for monitoring disease progression and as a biological outcome measure in tau-targeted therapeutic trials.

## Introduction

Progressive supranuclear palsy (PSP) is a neurodegenerative disease characterized by vertical supranuclear gaze palsy, postural instability, and cognitive decline.^1^ Neuropathologically, PSP is a primary four-repeat (4R) tauopathy featuring selective accumulation of straight tau filaments, distinguishing it from the paired helical filaments observed in 3R/4R tauopathies, such as Alzheimer’s disease.^2, 3^ Abnormal tau aggregates affect the neurons and glia, with tufted astrocytes and globose neurofibrillary tangles serving as hallmark features.^2^ These tau pathologies presumably originate in the pallido–nigro–luysian axis and subsequently propagate to the striatum, cerebellar dentate nucleus, and frontoparietal cortices.^4, 5^

Tau positron emission tomography (PET) enables *in vivo* visualization and quantification of tau pathologies.^6^ However, reliable imaging of 4R tau aggregates has long been considered challenging.^7^ First-generation tracers have exhibited insufficient utility in PSP owing to low 4R tau affinity and off-target binding.^7-10^ To improve 4R tau detection, second-generation tracers have been developed; however, many remain structural analogs of the first-generation tracer flortaucipir (18F) and enable only modest improvements in 4R tau pathology detection.^11, 12^ For example, [^18^F]PI-2620 has demonstrated encouraging diagnostic performance in PSP; however, neuropathological and imaging studies indicate that tracer uptake may vary according to the underlying composition of 4R tau pathology, warranting careful consideration in the quantitative assessment and biological interpretation of tau burden.^13-15^

To address the abovementioned limitations, florzolotau (18F) ([^18^F]PM-PBB3; [^18^F]APN-1607; hereafter florzolotau) was developed through enhanced binding affinity across diverse tau deposits.^16^ Cross-sectional studies have shown disease-specific tau distribution patterns in PSP with high diagnostic accuracy and demonstrated that regional tau burden correlates with disease severity.^17, 18^ Furthermore, a recent clinicopathological investigations confirmed a strong correspondence between *in vivo* PET signals and post-mortem tau burden in the same individuals.^19^

However, longitudinal validation of tau PET for tracking disease progression remains limited. Previous longitudinal studies using flortaucipir (18F) and [^18^F]PI-2620 detected increased uptake in the basal ganglia and cortical regions but failed to establish meaningful correlations with clinical decline.^20, 21^ These limitations highlight the need for longitudinal evaluation using florzolotau to determine whether it can reliably capture *in vivo* tau accumulation and its association with clinical progression in PSP.

Such an evaluation is crucial for therapeutic development. Numerous disease-modifying therapy (DMTs) trials in PSP have been conducted, most of which rely on clinical progression markers such as the PSP Rating Scale scores as primary endpoints to evaluate treatment efficacy.^22-25^ However, these symptom-based measures are subjective and variable and do not directly reflect pathological burden or target engagement. Thus, there is an urgent unmet need for objective biomarkers capable of quantifying longitudinal tau accumulation and monitoring therapeutic responses.^26, 27^

In the evaluation of florzolotau PET as a longitudinal tau imaging biomarker, analytical approaches must account for two key challenges: the widespread, multiregional nature of PSP pathology and the strong inter-regional covariance inherent in PET data. Conventional univariate methods used to examine individual regions may fail to capture spatial patterns of tau accumulation and lack statistical stability when multiple correlated regions are analyzed, which is particularly problematic in rare disorders like PSP with limited sample sizes. Partial least squares (PLS) analysis addresses these issues by identifying latent components that capture shared variance across the brain, enabling the identification of spatial patterns associated with group differences or clinical measures.^28, 29^ This multivariate approach has been increasingly used in neuroimaging studies to identify disease-related patterns that may be overlooked by conventional univariate analyses.^30-32^

This study aimed to investigate the spatiotemporal tau accumulation in PSP using one-year longitudinal florzolotau PET and to assess its utility as an imaging biomarker for tracking disease progression. Through PLS analysis, three aspects of tau pathology in PSP were characterized: (1) baseline distribution, (2) longitudinal accumulation, and (3) longitudinal changes associated with clinical progression.

## Methods

### Participants and Clinical Assessments

A total of 26 patients with PSP were recruited from affiliated hospitals, including 18 with probable PSP Richardson syndrome (PSP-RS) and 8 with PSP-non-RS (7 with probable PSP-parkinsonism [PSP-P] and 1 with probable PSP-frontal presentation [PSP-F]). PSP diagnosis was based on the International Parkinson and Movement Disorder Society clinical diagnostic criteria for PSP.^1^ Furthermore, 10 HCs without history of neurological or psychiatric disorders were recruited from the volunteer association of our institute. For longitudinal evaluation, the participants underwent neuropsychological assessments, florzolotau PET, and MRI scans at baseline and after 1 year. All participants, except for one with PSP, underwent [^11^C]Pittsburgh Compound B ([^11^C]PiB) PET within 3 months before the baseline assessment, and Aβ negativity was confirmed by visual inspection performed by at least three experienced PET specialists.^33^

Written informed consent, in accordance with the Declaration of Helsinki, was obtained from all participants and, if the participants were cognitively impaired, spouses or other close family members. This study was approved by the Radiation Drug Safety Committee and National Institutes for Quantum Science and Technology Certified Review Board of Japan. The study was registered with the UMIN Clinical Trials Registry (number 000030248).

### Image Acquisition and Data Preprocessing

#### Magnetic resonance imaging studies

MR images were acquired using a 3-T MAGNETOM Verio scanner (Siemens Healthcare). Three-dimensional T1-weighted gradient-echo sequence images were obtained for PET image co-registration and tissue segmentation (sagittal orientation; TE = 1.95 ms; TR = 2,300 ms; TI = 900 ms; flip angle = 9°; acquisition matrix = 512 × 512 × 176; voxel size = 1 × 0.488 × 0.488 mm).

#### PET studies

Florzolotau PET scans were performed using a Biograph mCT Flow system (Siemens Healthcare; matrix size 200 □× □ 200 □× □109; voxel size [mm] 2 □× □2 □× □2). Images were reconstructed using a filtered back-projection algorithm with a Hanning filter (4.0-mm full-width at half-maximum). Each participant underwent 90–110 min of PET scans (frames: 4 □× □ or 2 □× □ 10□min) following intravenous injection of florzolotau (average dose of 186.2 ± 8.0 MBq for baseline and 184.5 ± 7.3 MBq for follow-up). The PET scan protocols have been previously described,^16, 34^ and radiosynthesis procedures are detailed in the Supplementary Method.

The acquired PET images were corrected for head motion by aligning frames from 100–110 min to a summation image of 90–100 min using PMOD 4.3 and 4.4 (PMOD Technologies Ltd). Motion-corrected PET images were rigidly co-registered to individual T1-weighted MR images. Standardized uptake value ratio (SUVR) images were generated by averaging motion-corrected PET frames acquired 90–110 min postinjection and then dividing regional uptake by the mean activity of the reference region described below. Partial volume correction (PVC) was not applied in the primary analyses. To evaluate the potential impact of progressive atrophy on longitudinal SUVR changes, an additional validation analysis using the geometric transfer matrix method was conducted;^35^ the results are reported in the Supplementary Materials.

### Reference Region Definition

In 4R tauopathies, tau deposition in conventional regions, such as the cerebellar gray matter, may result in the underestimation of tracer binding in target regions.^9, 36^ To reduce this potential bias, reference regions were automatically determined using a histogram-based algorithm implemented in MATLAB (The MathWorks, Natick, MA, USA) that identifies gray matter voxels with low tau accumulation based on signal intensity histograms, as previously described.^34^ To ensure longitudinal consistency, a unified reference region approach was employed rather than independently defining reference regions at each time point. Reference regions were defined on follow-up PET images, as voxels retaining low tracer uptake at later disease stages are expected to serve as a more stable reference for longitudinal quantification. Subsequently, the reference region was transformed back to the baseline space using transformation matrices derived from the co-registration of follow-up and baseline MR images (Fig. S1).

### Region of Interest Definition

Individual T1-weighted MR images were automatically parcellated using M-Vision brain (M Corporation, Tokyo, Japan) following the structural definitions of the Johns Hopkins University brain atlas.^17, 37^ Then, these anatomically defined regions of interest (ROIs) were applied to co-registered PET images. Of the 144 ROIs at the fourth hierarchical level, 114 that corresponded to the brain parenchyma were selected. As PSP typically exhibits minimal hemispheric asymmetry in tau accumulation, homologous left and right ROIs were averaged to enhance measurement stability, yielding 57 bilateral ROIs for subsequent analyses.

### Statistical Analysis

#### General statistical analysis

All statistical analyses were conducted using JMP Pro 18 (SAS Institute Inc., Cary, NC, USA). The normality of data distributions was evaluated using the Shapiro–Wilk test. Owing to the non-normal distribution of several key variables and the relatively small sample size, nonparametric tests were applied for group comparisons and correlation analyses. Group comparisons of demographic and clinical characteristics were performed using the Mann–Whitney U test and chi-square test for continuous and categorical variables, respectively. Longitudinal changes in clinical scores were evaluated using the Wilcoxon signed-rank test. The annual changes (Δ) were calculated as the difference between follow-up and baseline values divided by the scan interval in years. Statistical significance was set at *p* < 0.05 (two-tailed).

#### Partial least squares analysis

PLS analysis is a multivariate statistical approach that extracts latent components, maximizing the covariance between explanatory variables (*X*) and a response variable (*Y*), enabling the assessment of individual variable contributions.^38^ This framework includes PLS discriminant analysis (PLS-DA) to identify patterns discriminating categorical groups and PLS regression (PLS-R) to determine associations with continuous outcomes. Both approaches were applied across 57 ROIs to characterize three aspects of tau pathology in PSP:

1. Baseline tau distribution (baseline PLS-DA): Baseline regional SUVRs (*X*) discriminating PSP from HCs (*Y*).
2. Longitudinal tau accumulation (longitudinal PLS-DA): Regional ΔSUVRs (*X*) discriminating PSP from HCs (*Y*).
3. Clinical progression-related tau accumulation (PLS-R): Regional ΔSUVRs (*X*) associated with ΔPSP Rating Scale scores (*Y*). Analyses were conducted in the overall PSP cohort and separately in PSP-RS, the most common and clinically representative phenotype.

Before analysis, regional SUVR values were adjusted for age and sex using a linear regression model. For the PLS-DA, the adjusted values were standardized to z-scores based on the mean and standard deviation of the HC group. In the PLS-R analysis in the PSP group, all variables, including imaging variables and the ΔPSP Rating Scale scores, were mean-centered and scaled to unit variance. PLS model estimation was performed using the nonlinear iterative PLS algorithm. The optimal number of latent components was selected by minimizing the predicted residual sum of squares through leave-one-out cross-validation. Model stability was assessed using the cross-validated *Q²* statistic, with *Q²* of >0 considered to indicate a reliable model. To evaluate the potential for overfitting, a permutation test with 1,000 iterations was conducted by randomly permuting the outcome variable, and statistical significance was defined as *p* < 0.05. ROIs with variable importance in projection (VIP) scores > 1.5 and positive regression coefficients were considered primary contributors to the models.^32, 39, 40^ Further mathematical details of the PLS algorithm are available in the Supplementary Method.

#### ROI-level analyses

To further characterize the spatial patterns identified by the PLS models, complementary analyses were conducted on ROIs with VIP scores of >1.5. Longitudinal changes in regional SUVR were evaluated using the Wilcoxon signed-rank test. Associations between ΔSUVR and ΔPSP Rating Scale scores were evaluated using Spearman’s rank correlation. Furthermore, linear mixed-effects models were used to evaluate longitudinal associations, with SUVR as the dependent variable, PSP Rating Scale scores and scan timing (baseline vs. follow-up) as fixed effects, and subjects as a random effect to account for repeated measurements. As these analyses were conducted on regions derived from the same dataset, they were considered exploratory *post hoc* analyses and not corrected for multiple comparisons.

## Results

### Demographic and Clinical Characteristics

The participants’ demographic and clinical characteristics are summarized in Table 1. At baseline, the PSP-RS group exhibited higher baseline PSP Rating Scale scores than the non-RS group (median [interquartile range (IQR)] 36 [26–49] vs. 19 [16–34], *p* = 0.026). Furthermore, over the approximately 1-year follow-up period, the PSP-RS group exhibited a substantial decline in the PSP Rating Scale scores (median annual change [IQR]: 7.6 [4.5–15.5], *p* < 0.001), whereas the non-RS group showed no significant change (median annual change [IQR]: 3.3 [-2.3–10.9], *p* = 0.219). Individual longitudinal trajectories of the PSP Rating Scale scores from baseline to follow-up for each group are presented in Fig. S2.

**Table 1.**
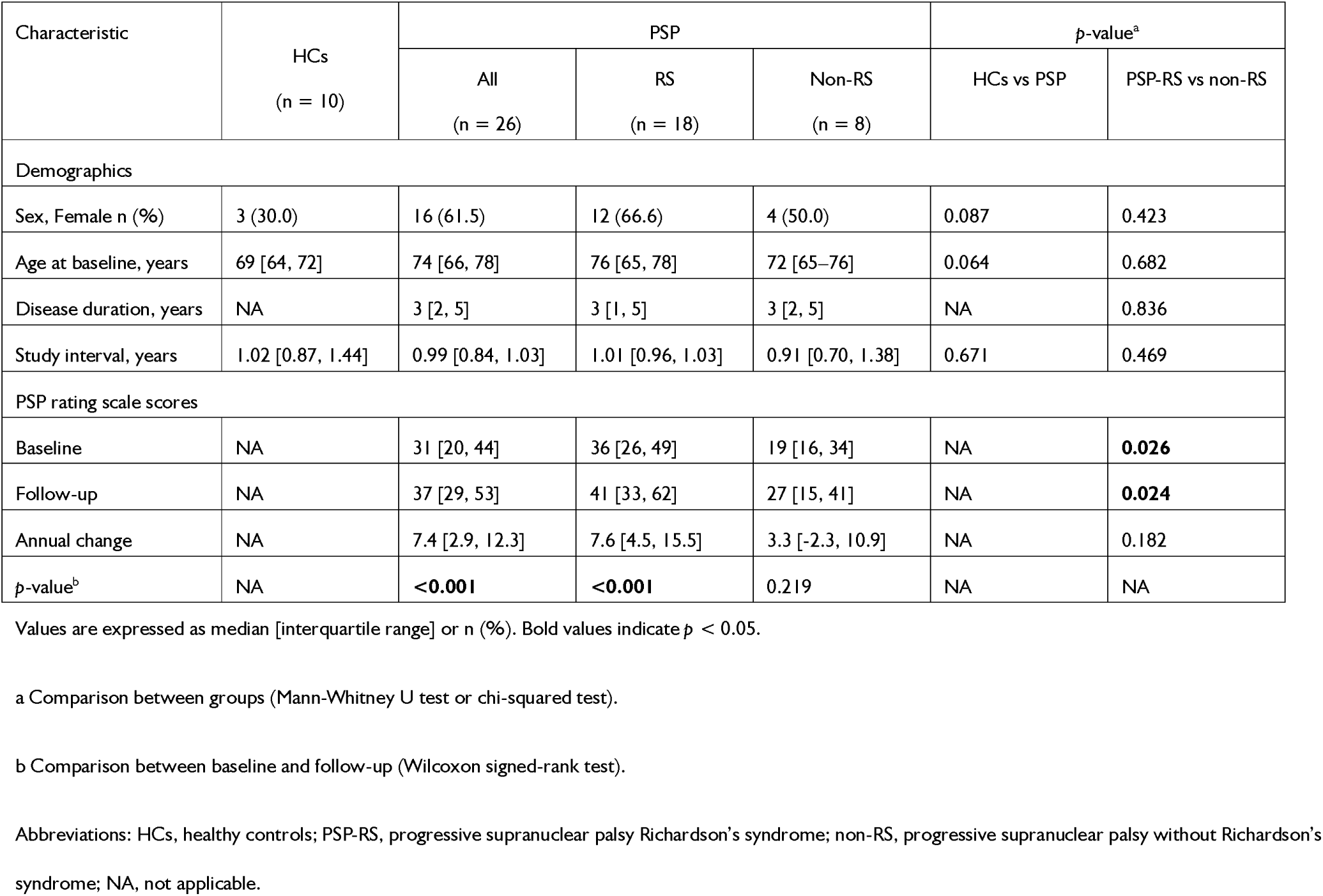
Demographic and clinical characteristics of the study participants.

| Characteristic | HCs<br>(n = 10) | PSP |  |  | p-value<sup>a</sup> |  |
| --- | --- | --- | --- | --- | --- | --- |
|  |  | All<br>(n = 26) | RS<br>(n = 18) | Non-RS<br>(n = 8) | HCs vs PSP | PSP-RS vs non-RS |
| Demographics |  |  |  |  |  |  |
| Sex, Female n (%) | 3 (30.0) | 16 (61.5) | 12 (66.6) | 4 (50.0) | 0.087 | 0.423 |
| Age at baseline, years | 69 [64, 72] | 74 [66, 78] | 76 [65, 78] | 72 [65–76] | 0.064 | 0.682 |
| Disease duration, years | NA | 3 [2, 5] | 3 [1, 5] | 3 [2, 5] | NA | 0.836 |
| Study interval, years | 1.02 [0.87, 1.44] | 0.99 [0.84, 1.03] | 1.01 [0.96, 1.03] | 0.91 [0.70, 1.38] | 0.671 | 0.469 |
| PSP rating scale scores |  |  |  |  |  |  |
| Baseline | NA | 31 [20, 44] | 36 [26, 49] | 19 [16, 34] | NA | **0.026** |
| Follow-up | NA | 37 [29, 53] | 41 [33, 62] | 27 [15, 41] | NA | **0.024** |
| Annual change | NA | 7.4 [2.9, 12.3] | 7.6 [4.5, 15.5] | 3.3 [-2.3, 10.9] | NA | 0.182 |
| p-value<sup>b</sup> | NA | **<0.001** | **<0.001** | 0.219 | NA | NA |

Values are expressed as median [interquartile range] or n (%). Bold values indicate $p < 0.05$ .
a Comparison between groups (Mann-Whitney U test or chi-squared test).
b Comparison between baseline and follow-up (Wilcoxon signed-rank test).
Abbreviations: HCs, healthy controls; PSP-RS, progressive supranuclear palsy Richardson's syndrome; non-RS, progressive supranuclear palsy without Richardson's syndrome; NA, not applicable.

### Baseline Tau Distribution and Longitudinal Tau Accumulation

The baseline PLS-DA identified a characteristic tau distribution that discriminated patients with PSP from HCs (Fig. 1A; see Table S1 for detailed model metrics). ROIs with primary contribution to this model were predominantly localized in subcortical regions along the pallido–nigro–luysian axis, particularly in the globus pallidus (GP) and midbrain (VIP = 2.62 and 1.94). High VIP scores were also observed in anatomically adjacent ROIs, including the nucleus accumbens and basal forebrain.

**Fig. 1.**
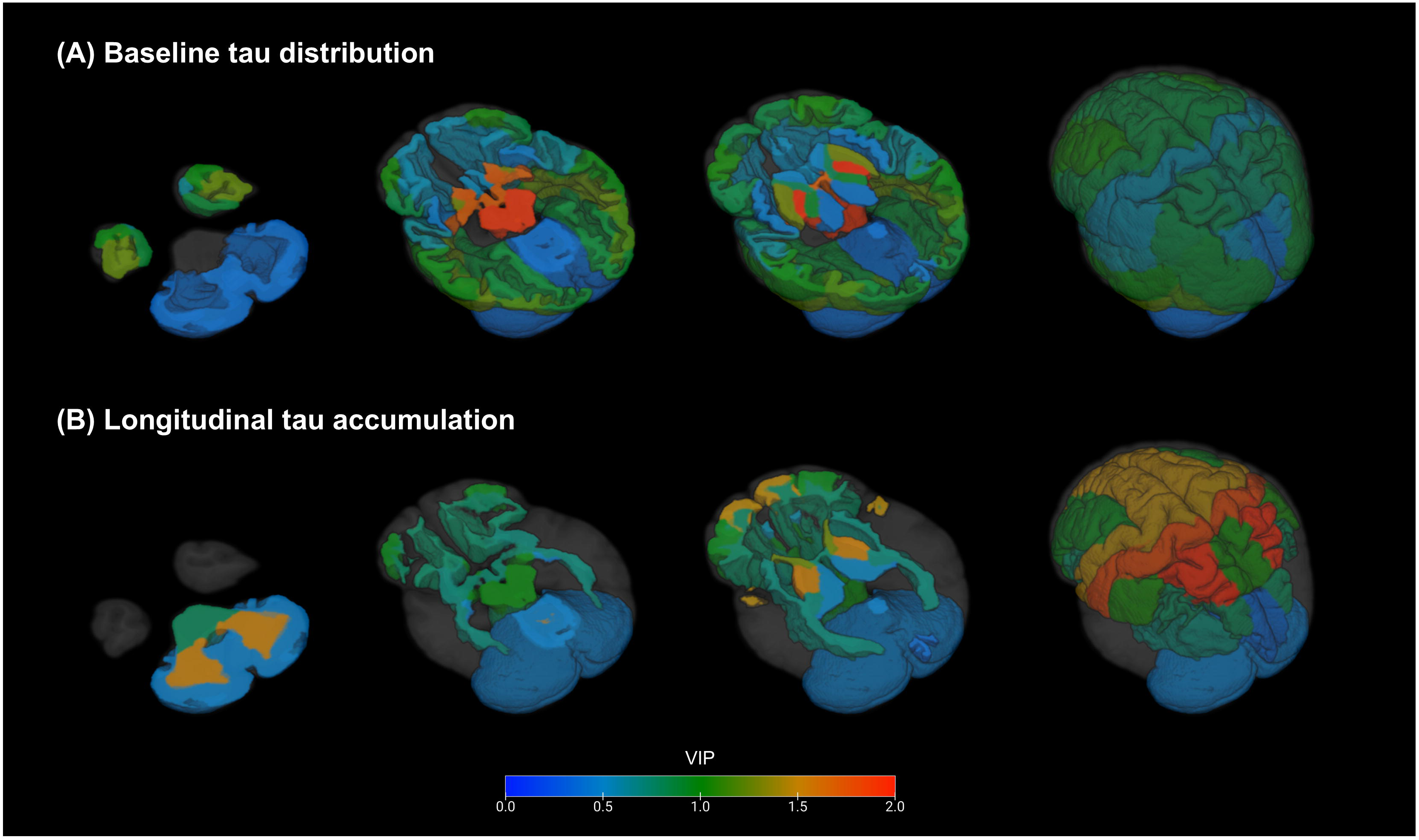
Variable Importance in Projection (VIP) scores map of (A) baseline and (B) longitudinal partial least squares discriminant analysis (PLS-DA) discerning patients with progressive supranuclear palsy (PSP) from healthy controls (HCs). Brain regions with positive coefficients, indicating higher tau accumulation in patients with PSP than in HCs, are displayed. Regions are color-coded according to their VIP scores, with warmer colors reflecting stronger contribution to group discrimination. Images show 3D-rendered axial sections at three anatomical levels (cerebellum, midbrain, and basal ganglia), followed by whole-brain surface renderings. In all panels, the left side of the image corresponds to the left hemisphere. Abbreviations: HCs, healthy controls; PSP, progressive supranuclear palsy; SUVR, standardized uptake value ratio; VIP, variable importance in projection.

Contrary to the cross-sectional distribution, the longitudinal PLS-DA revealed a distinct tau accumulation with a broader regional involvement (Fig. 1B; see Table S1 for detailed model metrics). Among the ROIs highlighted at baseline, the GP continued to demonstrate a high contribution (VIP = 1.57), whereas the midbrain showed a reduced contribution (VIP = 0.99). Beyond these subcortical regions, ROIs with primary contribution extended to the parietal cortex (e.g., superior parietal gyrus; VIP = 2.27) and cerebellar white matter (VIP = 1.56). Furthermore, in the frontal cortex, a region involved in PSP tau pathology, a few ROIs exhibited high VIP scores approaching the primary contributor threshold (e.g., superior frontal gyrus; VIP = 1.48). The posterior limb of the internal capsule, which is anatomically adjacent to the GP, also reached the predefined cutoff.

These patterns were supported by univariate analyses of key regions (Fig. 2). At baseline, patients with PSP exhibited higher SUVR than HCs in the GP (median 1.57 vs. 1.27, *p* < 0.001) and midbrain (1.51 vs. 1.24, *p* = 0.001), with an additional difference observed in the superior parietal gyrus (*p* = 0.009). Over the follow-up period, tau accumulation was observed in the superior parietal gyrus (median ΔSUVR = 0.030, representing a 3.3% annual increase relative to baseline; *p* = 0.009), GP (0.034, 2.2% annual increase; *p* = 0.005), and cerebellar white matter (0.029, 2.2% annual increase; *p* = 0.047), with direct group comparisons of ΔSUVR confirming accelerated accumulation in these regions compared with HCs. Contrarily, no significant increase was observed in the midbrain (0.004, 0.3% annual increase; *p* = 0.480). The receiver operating analyses for the differentiation of PSP from HCs using baseline SUVR and ΔSUVR are presented in Fig. S3, and detailed baseline and longitudinal data for all ROIs are provided in Table S2.

**Fig. 2.**
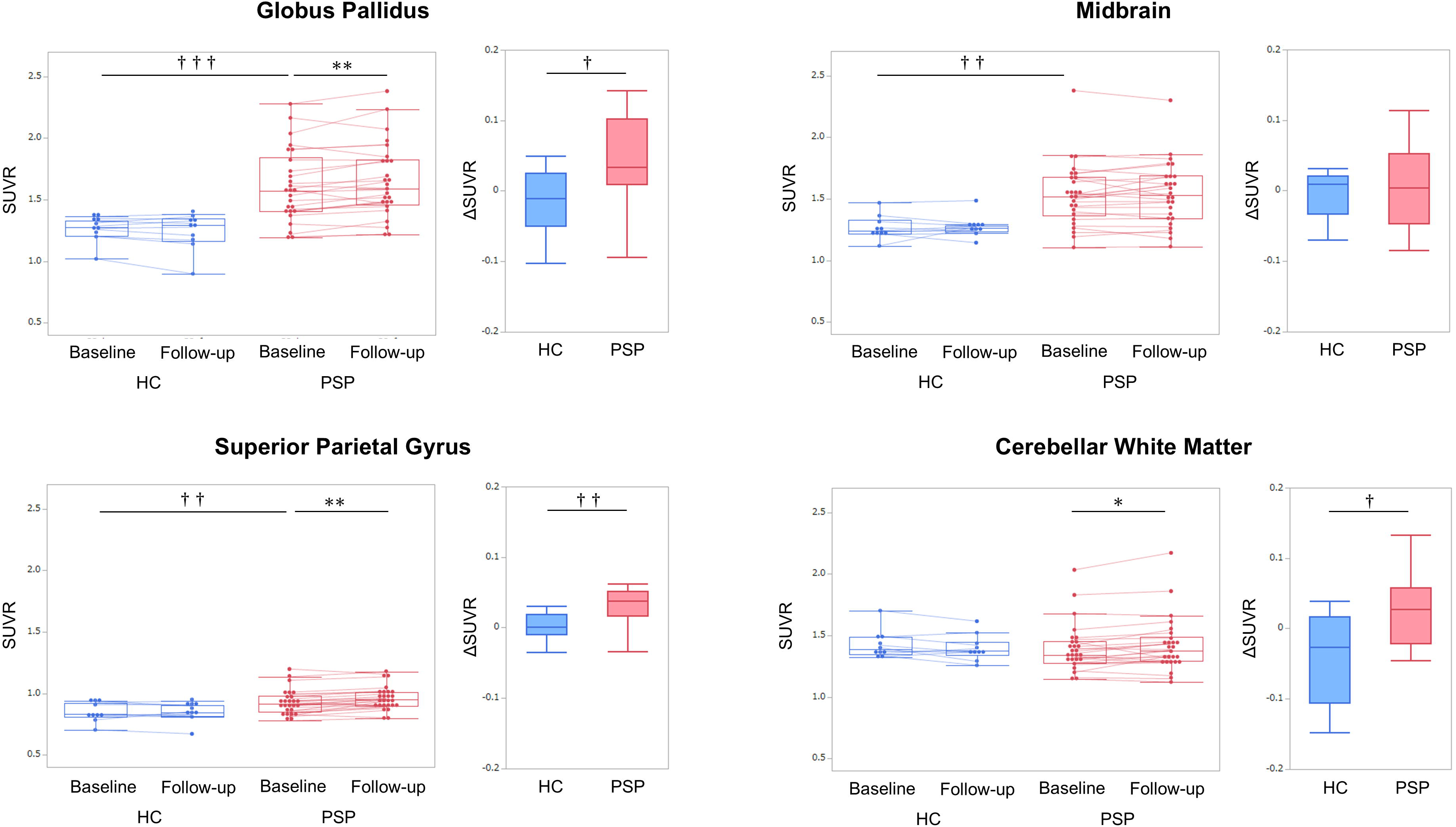
Pairs of graphs are displayed for four key regions with primary contribution (VIP score > 1.5) to the baseline or longitudinal PLS-DA model. For each region, comparison of standardized uptake value ratios between baseline and follow-up for healthy controls (HCs, blue) and patients with progressive supranuclear palsy (PSP, red) in the left graphs, with individual trajectories are connected by lines. Direct group comparisons of annualized ΔSUVR between the HC and PSP groups are presented in the right graphs. Asterisks denote significant longitudinal changes within each cohort (* p < 0.05, ** p < 0.01, *** p < 0.001). Daggers indicate significant differences between HCs and PSP in either baseline SUVR or ΔSUVR († p < 0.05, †† p < 0.01, ††† p < 0.001). Abbreviations: SUVR, standardized uptake value ratio; HCs, healthy controls; PLS-DA, partial least square discriminant analysis; PSP, progressive supranuclear palsy; VIP, variable importance in projection.

### Clinical Progression–Related Tau Accumulation

In the entire PSP cohort, the PLS-R model did not show a stable multivariate association (*Q^2^* < 0). However, when the analysis was restricted to the PSP-RS group, the model identified meaningful clinical progression–related regional tau accumulation (Figure 3A; see Table S1 for detailed model metrics). The GP showed the highest contribution (VIP = 2.06), followed by the pons (VIP = 1.56). In the univariate analyses, ΔSUVR in the GP exhibited a strong positive correlation with ΔPSP Rating Scale scores (Spearman’s ρ = 0.688, 95% confidence interval [CI]: 0.326 to 0.874, *p* = 0.002; Fig. 3B), whereas the pons showed a moderate correlation (Spearman’s ρ = 0.552, 95% CI: 0.115 to 0.810, *p* = 0.018).

**Fig. 3.**
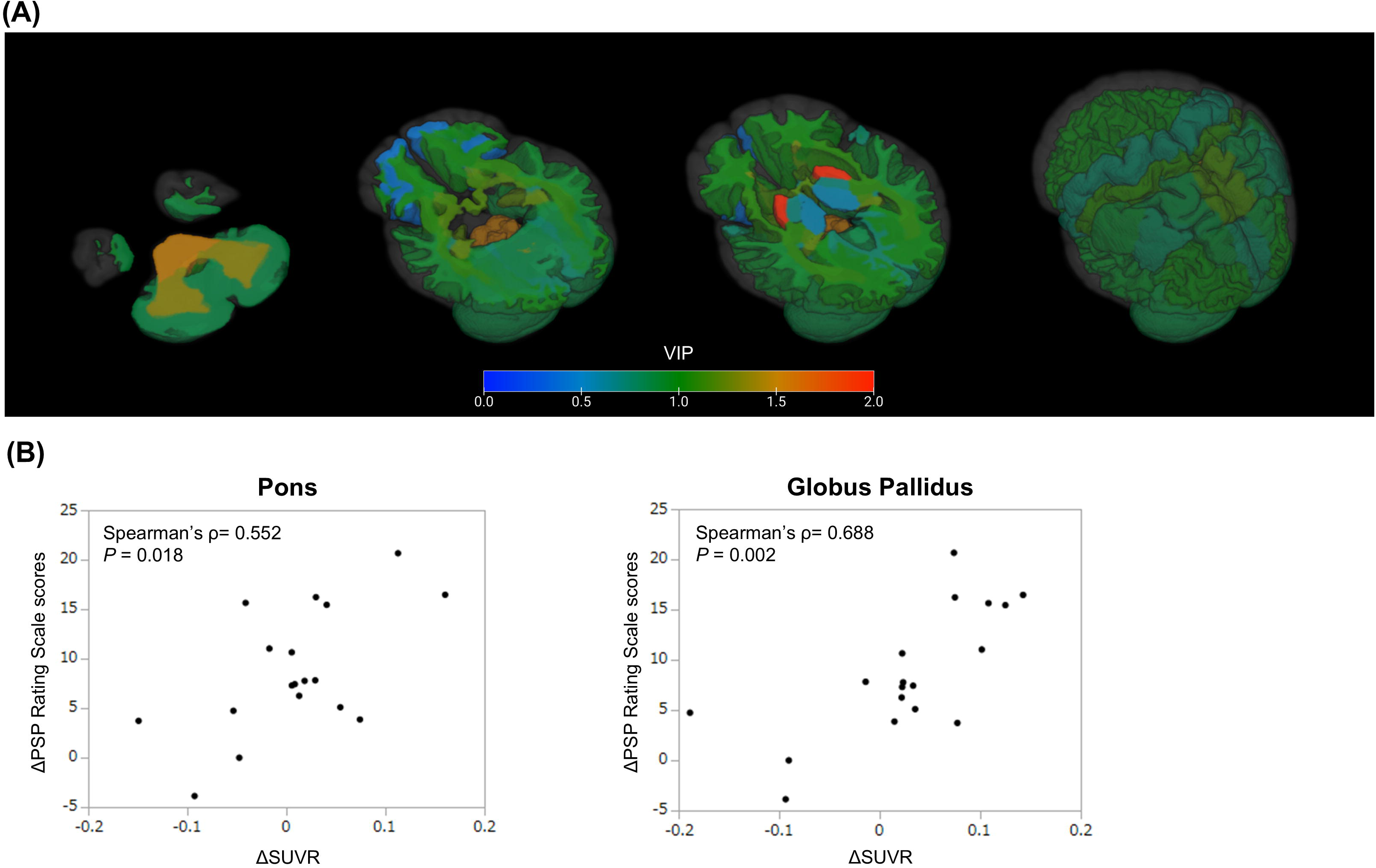
(A) Variable Importance in Projection (VIP) scores map of partial least squares regression (PLS-R) identifying regional tau accumulation (ΔSUVR) associated with clinical progression (ΔPSP Rating Scale scores) in PSP-RS. Regions with positive coefficients are color-coded according to their VIP scores, with warmer colors reflecting stronger contribution to the model. Images depict 3D-rendered axial sections at three anatomical levels (cerebellum, midbrain, and basal ganglia), followed by whole-brain surface renderings. In all panels, the left side of the image corresponds to the left hemisphere. (B) Scatter plot demonstrating the association between ΔSUVR and ΔPSP Rating Scale scores in PSP-RS. Data are presented for the globus pallidus and pons, key regions with a primary contribution (VIP score > 1.5) to the PLS-R model. Abbreviations: PLS-R, partial least squares regression analysis; PSP-RS, progressive supranuclear palsy Richardson’s syndrome; SUVR, standardized uptake value ratio; VIP, variable importance in projection.

Figure 4 presents individual SUVR trajectories plotted against PSP Rating Scale scores for the ROIs identified in the PLS analyses (VIP > 1.5). Among these regions, the GP uniquely showed cross-sectional correlation with disease severity at baseline (Spearman’s ρ = 0.398, 95% CI: 0.012 to 0.680, *p* = 0.044) and the strongest positive association in linear mixed-effects models (β = 0.0043, 95% CI: 0.0008 to 0.0078, *p* = 0.018), with consistently increasing trajectories across the disease spectrum. Complete statistics for the linear mixed-effects models across all investigated regions are summarized in Table S3.

**Fig. 4.**
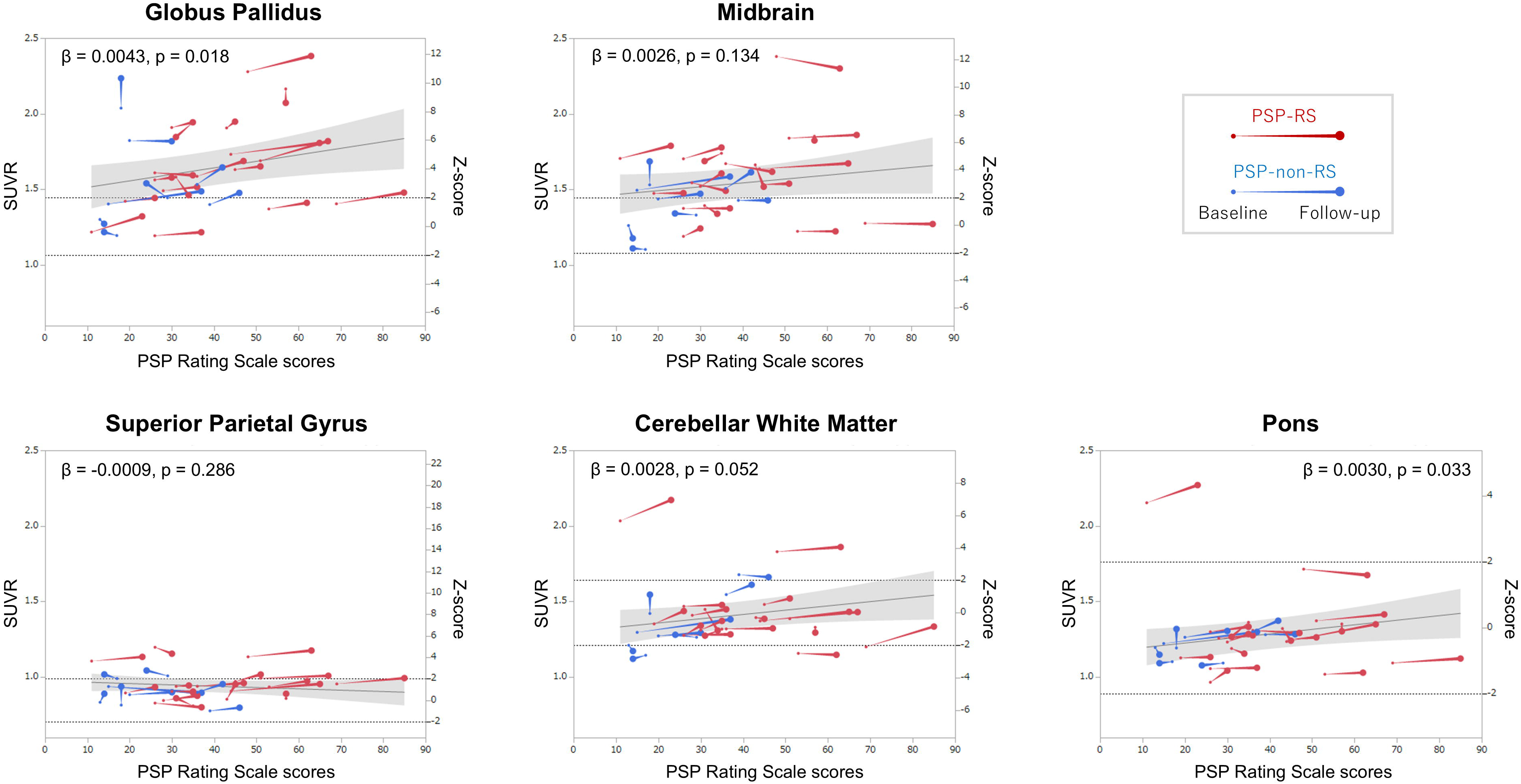
Individual trajectories of regional SUVRs in relation to PSP Rating Scale scores. Regions with variable importance in projection (VIP) > 1.5 identified across PLS analyses are presented: globus pallidus, midbrain, superior parietal gyrus, cerebellar white matter, and pons. Each line denotes the trajectory of an individual participant from baseline to follow-up, illustrating within-subject longitudinal changes. Red lines denote patients with PSP-RS, whereas blue lines represent those with PSP-non-RS. Smaller dots indicate baseline values, whereas larger dots denote follow-up values. Z-scores represent deviations from the baseline mean of healthy controls. Horizontal dashed lines indicate Z = ±2, representing thresholds commonly used to indicate abnormal deviation from the healthy control range. The regression line represents the fixed effect of the PSP Rating Scale score estimated by a linear mixed model. Shaded areas indicate 95% confidence intervals of the regression lines. β coefficients and p values for the PSP Rating Scale score from the models are presented in each panel. Abbreviations: PLS, partial least squares; PSP-RS, progressive supranuclear palsy Richardson’s syndrome; SUVR, standardized uptake value ratio; VIP, variable importance in projection.

## Discussion

This longitudinal florzolotau PET study examined spatiotemporal changes in tau accumulation in PSP and their relationship to clinical progression. Contrary to subcortical-predominant distributions at baseline, longitudinal analyses over a 1-year period revealed a more extensive accumulation involving the neocortical and cerebellar regions. A key finding was the prominent involvement of the GP, which was consistently observed in the baseline and longitudinal analyses. Essentially, longitudinal increases in florzolotau uptake in the GP exhibited a positive correlation with clinical progression in the PSP-RS group. To our knowledge, this is the first study to demonstrate such a longitudinal relationship *in vivo*, extending the potential utility of florzolotau PET as an objective imaging biomarker from diagnostic assessment to disease progression monitoring in PSP.

In the baseline cross-sectional analysis, florzolotau uptake was predominantly localized to the pallido–nigro–luysian axis, particularly the GP and midbrain, consistent with previous tau PET studies.^16-19^ This pattern aligns with established neuropathological staging models, which identify these subcortical structures as the earliest sites of dense 4R tau deposition in neurons and glia.^5, 41^ These findings suggest that the baseline tau distribution pattern in this study reflects a highly discriminative spatial signature of the core pathological features of PSP.

Beyond these baseline observations, the longitudinal analysis revealed distinct florzolotau accumulation across more widespread regions, including the GP, frontoparietal cortex, and cerebellar white matter. This spatial pattern is consistent with the neuropathological staging models of PSP, which suggest the delayed emergence of cortical and cerebellar tau deposition in the disease course, following initial subcortical involvement.^5^ Although such hierarchical spreading of tau pathology has been previously modeled by cross-sectional tau PET studies,^18, 42^ the longitudinal tau accumulation observed in this study provides *in vivo* evidence of spatiotemporal progression in PSP. These findings indicate the sensitivity of florzolotau PET in detecting meaningful tau-related changes over a 1-year period.

Consistent with neuropathological evidence indicating that the GP is a core region of tau pathology in PSP,^4^ this region exhibited prominent florzolotau uptake in the baseline and longitudinal analyses. Notably, in PSP-RS, the most common and representative PSP phenotype, longitudinal tau accumulation in this region was most strongly associated with clinical progression, indicating the highest multivariate importance and significant univariate correlation. Extending its previously established role as a diagnostic biomarker,^17, 18^ these findings highlight the use of florzolotau PET for longitudinal disease monitoring and suggest its potential as an outcome measure for future DMTs targeting tau pathology.

Contrarily, the midbrain demonstrated prominent florzolotau uptake at baseline, whereas longitudinal increases were relatively limited. This observation may reflect the advanced stage of neurodegeneration in this region, where substantial neuronal loss has already occurred.^20, 43^ In such contexts, the clearance of tau aggregates due to neuronal loss may counterbalance continued tau accumulation in surviving neurons and glia, resulting in an apparent plateau in tracer uptake. As a preliminary validation, the potential impact of atrophy progression on longitudinal SUVR changes was assessed by comparing data with and without PVC. The midbrain continued to demonstrate no longitudinal change following PVC, whereas other regions showed patterns consistent with the uncorrected analysis (Table S4), suggesting that atrophy-related partial volume effects are unlikely to fully account for the observed plateau.

The lack of a consistent association between longitudinal florzolotau accumulation and clinical progression across the entire PSP cohort highlights the clinical heterogeneity of the disease. Several factors may account for this finding. First, although non-RS subtypes may converge toward PSP-RS–like phenotypes, they are characterized by greater heterogeneity in clinical trajectories and spatial patterns of tau deposition.^44, 45^ Second, previous studies have consistently reported a slower rate of clinical progression in non-RS variants than PSP-RS.^45-48^ Consistent with these observations, patients with non-RS subtypes in the present study exhibited smaller changes in the PSP Rating Scale over the follow-up period. Slow clinical progression and pronounced phenotypic heterogeneity in non-RS subtypes may have reduced statistical power to detect meaningful longitudinal associations.

As an alternative to the histogram-based gray matter reference used in this study (GM-ref; Fig. S4A), white matter references have been adopted in some tau PET studies owing to their minimal 4R tau burden and reduced variability compared with conventional cerebellar references.^49, 50^ Preliminary analyses using a histogram-based white matter reference (WM-ref; Fig. S4B) largely preserved baseline discrimination between PSP and HCs and longitudinal changes within PSP (Fig. S5). However, the WM-ref failed to demonstrate group differences in ΔSUVR or detect the correlation between GP tau accumulation and clinical progression in PSP-RS observed with the GM-ref (Fig. S5 and Fig. S6). The lack of group differences in ΔSUVR may reflect age-related changes in white matter tracer retention, which have been reported to introduce systematic bias into longitudinal measurements.^34^ The reduced sensitivity to detect clinical correlations may, in turn, reflect increased variance arising from tissue-type mismatch between gray and white matter target and reference regions (Fig. S7), likely due to differences in tracer retention and nonspecific binding.^51^ Consistent with this interpretation, a previous longitudinal [^18^F]PI-2620 study using white matter reference regions failed to detect significant clinical correlations.^21^ These findings highlight the importance of reference region selection for longitudinal tau PET assessments. Additional comparative analyses and discussion of reference region strategies are provided in the Supplementary Discussion.

This study has several limitations. First, the sample size was relatively small, which may have affected the statistical power of identifying disease-related regions and limited phenotype-specific analyses. Second, as the present cohort mainly consisted of patients with mild-to-moderate disease severity, our findings may not be fully generalizable to patients with prodromal or advanced disease. Third, a single 1-year follow-up duration is a relatively limited observation window for a neurodegenerative disease and may not capture the nonlinear trajectories of tau accumulation. Fourth, although our image processing pipeline is data-driven and theoretically applicable to independent cohorts or other 4R tau PET tracers, empirical validation in external datasets is required to fully confirm its generalizability. Finally, multivariate PLS analysis was conducted in a relatively small cohort with a large number of explanatory variables. Although permutation testing supported the robustness of the findings, external validation is warranted to confirm their stability and generalizability, specificcally because demographic adjustments and z-score normalization were based on the study cohort.

In conclusion, this longitudinal florzolotau PET study provides *in vivo* evidence of the spatiotemporal progression of tau pathology in PSP, marked by progression from subcortical regions to the frontoparietal cortex and cerebellum. Tau accumulation in the GP emerges early and continues to increase alongside clinical deterioration in PSP-RS. These findings extend the clinical utility of florzolotau PET as an objective imaging biomarker, extending its role from diagnostic assessment to disease progression monitoring and serving as a biological outcome measure in future tau-targeted therapeutic trials.

## Acknowledgements

The authors thank all patients and their caregivers for their participation in this study, as well as clinical research coordinators, PET and MRI operators, radiochemists, and research ethics advisers at QST for their assistance with the current projects. We thank APRINOIA Therapeutics for kindly sharing a precursor of florzolotau. The authors acknowledge the support with patient recruitment and partial study implementation from: T. Takeda and I. Isose at Chiba-East Hospital; Masaki Oya at Funabashi Municipal Medical Center; Y. Takado at the Institute for Quantum Life Science, QST; T. Hatano, T. Tsunemi, N. Nishikawa, K. Nishioka, Y. Yamashita, Y. Motoi, and S. Saiki at Juntendo University; T. Yuasa at Kamagaya General Hospital; M. Seki at Keio University; S. Furukawa at Narita Red Cross Hospital; I. Aiba at National Hospital Organization Higashinagoya National Hospital; Y. Nishida and Y. Yagi at Tokyo Medical and Dental University (now Tokyo Science University); and H. Imai at Tokyo Rinkai Hospital.

## Author Roles

Y.C., K. Tagai, M.H., and H.E. contributed to drafting and revision of the manuscript for content, including medical writing; Y.C., K. Tagai, Y.K., R.G., K.O., H.M., M.I., S.M., Y.M., T.K., C.S., K.M., K.H., S.K., S.M., Y.K, H.K., Y.Y, Y.N., S.H., and K. Takahata contributed to the acquisition of data; Y.C., K. Tagai, H. Shinotoh, H. Shimada, M.H., and H.E. contributed to the study concept and design; Y.C., K. Tagai, A.O., C.S., K.K., M.-R.Z., T.T., M.H., and H.E. contributed to the analysis and interpretation of data.

## Data availability

Data supporting the findings of this study are available from the corresponding author upon reasonable request. Sharing and reuse of data require the expressed written permission of the authors, as well as clearance from the Institutional Review Boards.

