## Supplementary Materials for "Spatiotemporal Tau Accumulation and Its Clinical Impact on PSP: Longitudinal Florzolotau (18F) PET"

**Table S1.** Metrics and regional contributions of PLS models

|  | **Baseline PLS-DA** | **Longitudinal PLS-DA** | **PLS-R (PSP-RS)** |
| --- | --- | --- | --- |
| Predictor variable (X) | Baseline SUVR | ΔSUVR | ΔSUVR |
| Response variable (Y) | Group (PSP vs HC) | Group (PSP vs HC) | ΔPSP rating scale |
| Number of Latent components | 3 | 3 | 3 |
| R^2^Y | 0.658 | 0.515 | 0.840 |
| Q^2^ | 0.905 | 0.644 | 0.170 |
| Permutation *p*-value | < 0.001 | < 0.001 | 0.032 |
| **Regions of interest** | **VIP / Coefficient** | | |
| Superior Frontal Gyrus | 0.87 / 0.005 | 1.48 / 0.022 | 1.00 / -0.077 |
| Middle Frontal Gyrus | 1.02 / 0.009 | 0.96 / 0.007 | 1.15 / -0.086 |
| Inferior Frontal Gyrus | 0.85 / 0.008 | 1.14 / -0.025 | 0.86 / -0.089 |
| Occipital Gyrus | 0.57 / 0.003 | 0.99 / -0.016 | 0.37 / 0.019 |
| Rectal Gyrus | 0.42 / -0.002 | 1.52 / -0.068 | 1.08 / -0.093 |
| Postcentral Gyrus | 0.59 / 0.008 | **1.85 / 0.034** | 0.55 / -0.014 |
| Precentral Gyrus | 0.87 / 0.016 | 1.47 / 0.038 | 0.76 / 0.072 |
| Superior Parietal Gyrus | 0.83 / 0.014 | **2.27 / 0.042** | 0.87 / 0.050 |
| Supramarginal Gyrus | 0.60 / 0.009 | 1.05 / 0.004 | 0.92 / 0.021 |
| Angular Gyrus | 0.85 / 0.014 | 0.91 / -0.017 | 0.80 / 0.010 |
| Precuneus | 0.76 / 0.009 | 1.03 / 0.036 | 1.16 / 0.112 |
| Superior Temporal Gyrus | 0.65 / 0.006 | 1.24 / -0.036 | 0.87 / -0.051 |
| Middle Temporal Gyrus | 0.99 / 0.019 | 1.23 / -0.023 | 0.82 / -0.066 |
| Inferior Temporal Gyrus | 1.23 / 0.028 | 0.88 / -0.019 | 0.86 / -0.077 |
| Limbic Cortex | 0.47 / -0.010 | 0.73 / -0.022 | 1.34 / -0.122 |
| Fusiform Gyrus | 0.89 / 0.022 | 1.00 / -0.025 | 0.88 / 0.010 |
| Superior Occipital Gyrus | 0.56 / 0.006 | 0.81 / 0.016 | 0.91 / 0.045 |
| Middle Occipital Gyrus | 0.91 / 0.019 | 0.57 / -0.029 | 0.94 / -0.051 |
| Inferior Occipital Gyrus | 1.14 / 0.028 | 0.73 / -0.032 | 0.55 / -0.058 |
| Cuneus | 0.40 / 0.004 | 0.34 / 0.008 | 0.80 / 0.039 |
| Lingual Gyrus | 0.28 / -0.001 | 0.46 / -0.012 | 0.77 / 0.051 |
| Cingulate Gyrus | 0.53 / 0.000 | 1.62 / -0.068 | 0.87 / -0.069 |
| Insular Cortex | 0.43 / 0.000 | 1.01 / -0.031 | 0.60 / -0.033 |
| Amygdala | 0.48 / -0.002 | 0.99 / 0.029 | 0.31 / -0.006 |
| Hippocampus | 0.29 / -0.006 | 0.55 / -0.011 | 1.21 / 0.087 |
| Caudate Nucleus | 0.47 / 0.003 | 1.16 / -0.011 | 0.99 / -0.062 |
| Putamen | 1.28 / 0.019 | 0.86 / -0.006 | 0.76 / -0.010 |
| Globus Pallidus | **2.62 / 0.048** | **1.57 / 0.061** | **2.06 / 0.197** |
| Thalamus | 0.39 / 0.008 | 0.50 / 0.016 | 0.55 / 0.018 |
| Basal Forebrain | **1.743 / 0.028** | 0.75 / 0.004 | 1.18 / 0.061 |
| Nucleus Accumbens | **1.77 / 0.033** | 0.94 / -0.006 | 1.14 / 0.096 |
| Midbrain | **1.94 / 0.041** | 0.99 / 0.028 | 1.11 / -0.011 |
| Cerebellum Gray Matter | 0.36 / 0.012 | 0.45 / 0.019 | 0.85 / 0.086 |
| Pons | 0.32 / -0.008 | 0.81 / 0.036 | **1.56 / 0.112** |
| Medulla | 0.33 / -0.011 | 0.63 / 0.024 | 1.02 / 0.014 |
| Anterior Deep White Matter | 0.87 / -0.027 | 0.48 / -0.008 | 1.00 / 0.019 |
| Posterior Deep White Matter | 0.70 / -0.022 | 0.41 / 0.001 | 1.33 / 0.077 |
| Genu of the Corpus Callosum | 0.99 / -0.030 | 0.73 / -0.032 | 0.86 / -0.028 |
| Body of the Corpus Callosum | 0.40 / -0.011 | 0.68 / -0.002 | 1.14 / 0.058 |
| Splenium of the Corpus Callosum | 0.96 / -0.030 | 0.41 / -0.003 | 1.44 / 0.087 |
| Periventricular White Matter (lateral) | 0.31 / -0.008 | 0.38 / -0.013 | 1.05 / 0.097 |
| Anterior Limb of the Internal Capsule | 0.82 / 0.007 | 1.24 / 0.025 | 0.66 / -0.013 |
| Posterior Limb of the Internal Capsule | 0.92 / 0.002 | **1.57 / 0.033** | 0.72 / 0.016 |
| Inferior Deep White Matter | 1.18 / -0.039 | 0.72 / 0.006 | 1.07 / 0.068 |
| Cingulum (Cingulate Gyrus segment) | 0.76 / -0.016 | 0.59 / 0.013 | 1.00 / 0.002 |
| Cingulum (Hippocampal Segment) | 0.53 / -0.014 | 0.89 / -0.041 | 0.86 / -0.013 |
| Fornix / Stria Terminalis | 0.41 / -0.011 | 0.50 / 0.022 | 1.14 / 0.059 |
| Fornix | 0.47 / -0.010 | 0.22 / -0.010 | 0.69 / 0.020 |
| Peripheral Parietal White Matter | 1.14 / -0.011 | 0.81 / 0.021 | 1.08 / 0.057 |
| Periventricular White Matter (anterior) | 0.66 / -0.020 | 0.62 / -0.026 | 1.25 / 0.048 |
| Periventricular White Matter (posterior) | 0.47 / -0.012 | 0.57 / 0.020 | 1.01 / 0.077 |
| Peripheral Frontal White Matter | 3.29 / -0.010 | 0.78 / 0.004 | 0.98 / 0.005 |
| Peripheral Temporal White Matter | 1.14 / -0.012 | 1.05 / -0.023 | 0.93 / 0.003 |
| Peripheral Occipital White Matter | 0.58 / -0.001 | 0.84 / -0.008 | 0.99 / 0.070 |
| Peripheral Cingulate White Matter | 1.24 / -0.025 | 0.95 / 0.013 | 0.89 / -0.014 |
| Cerebellum White Matter | 0.53 / -0.016 | **1.56 / 0.075** | 1.35 / 0.082 |
| Caudate tail | 0.35 / -0.009 | 0.40 / 0.008 | 0.74 / 0.018 |

Data were derived from the entire PSP group and healthy controls for baseline and longitudinal PLS-DA, and from the PSP-RS group for PLS-R. R^2^Y denotes the cumulative percentage of variation in the response variable explained by the model. Q^2^ indicates the cumulative predictive relevance determined by cross-validation, where a positive value demonstrates the model’s reliability. The permutation *p*-value, calculated using 1,000 iterations, assesses the statistical significance of the model to ensure that the predictive performance is not due to overfitting. Bold values indicate Variable Importance in Projection (VIP) scores > 1.5 with positive coefficients, indicating higher SUVR or greater increase in PSP compared with controls.

Abbreviations: HC, healthy controls; PLS-DA, partial least squares discriminant analysis; PLS-R, partial least squares regression analysis; PSP, progressive supranuclear palsy; PSP-RS, progressive supranuclear palsy Richardson’s syndrome; SUVR, standardized uptake value ratio; VIP, variable importance in projection.

**Table S2.** Baseline SUVR and longitudinal changes across all regions

| **Regions of interest** | **Baseline SUVR** | | **ΔSUVR** | |
| --- | --- | --- | --- | --- |
|  | **HC** | **PSP** | **HC** | **PSP** |
| Superior Frontal Gyrus | 0.86 [0.81, 0.90] | 0.89 [0.86, 0.97] | 0.002 [-0.026, 0.016] | 0.018 [0.005, 0.046] |
| Middle Frontal Gyrus | 0.86 [0.85, 0.88] | 0.93 [0.87, 0.97] | 0.004 [-0.021, 0.009] | 0.011 [-0.008, 0.029] |
| Inferior Frontal Gyrus | 0.90 [0.85, 0.92] | 0.95 [0.88, 0.98] | 0.014 [-0.012, 0.023] | -0.002 [-0.022, 0.020] |
| Occipital Gyrus | 0.94 [0.88, 0.98] | 0.93 [0.91, 0.99] | 0.008 [-0.015, 0.033] | -0.020 [-0.047, 0.037] |
| Rectal Gyrus | 0.94 [0.90, 1.00] | 0.94 [0.88, 0.99] | 0.011 [-0.027, 0.021] | -0.009 [-0.024, 0.020] |
| Postcentral Gyrus | 0.83 [0.80, 0.88] | 0.87 [0.84, 0.94] | 0.005 [-0.007, 0.010] | 0.017 [0.001, 0.044] |
| Precentral Gyrus | 0.86 [0.85, 0.93] | 0.96 [0.89, 1.03] | 0.006 [-0.020, 0.019] | 0.028 [0.003, 0.043] |
| Superior Parietal Gyrus | 0.83 [0.81, 0.92] | 0.92 [0.85, 0.98] | -0.007 [-0.019, 0.010] | 0.030 [0.014, 0.047] |
| Supramarginal Gyrus | 0.89 [0.86, 0.97] | 0.93 [0.90, 0.95] | 0.001 [-0.011, 0.010] | 0.008 [-0.007, 0.029] |
| Angular Gyrus | 0.88 [0.86, 0.93] | 0.95 [0.93, 0.98] | 0.002 [-0.017, 0.020] | 0.020 [-0.006, 0.037] |
| Precuneus | 0.90 [0.87, 0.99] | 0.97 [0.93, 1.03] | 0.006 [-0.018, 0.027] | 0.032 [0.008, 0.054] |
| Superior Temporal Gyrus | 0.89 [0.87, 0.92] | 0.91 [0.86, 0.94] | -0.009 [-0.042, 0.004] | 0.001 [-0.014, 0.025] |
| Middle Temporal Gyrus | 0.95 [0.94, 0.98] | 1.00 [0.96, 1.03] | 0.002 [-0.013, 0.013] | -0.005 [-0.017, 0.020] |
| Inferior Temporal Gyrus | 0.98 [0.93, 1.03] | 1.03 [0.97, 1.08] | 0.010 [-0.020, 0.031] | -0.016 [-0.039, 0.021] |
| Limbic Cortex | 0.92 [0.87, 0.99] | 0.90 [0.86, 0.97] | -0.013 [-0.029, 0.034] | 0.011 [-0.022, 0.022] |
| Fusiform Gyrus | 1.04 [0.98, 1.10] | 1.05 [1.03, 1.11] | 0.015 [-0.008, 0.049] | -0.012 [-0.030, 0.017] |
| Superior Occipital Gyrus | 0.94 [0.91, 0.97] | 0.97 [0.93, 1.01] | -0.022 [-0.038, 0.011] | 0.022 [-0.008, 0.036] |
| Middle Occipital Gyrus | 0.94 [0.92, 0.96] | 0.99 [0.97, 1.04] | -0.001 [-0.016, 0.021] | 0.006 [-0.012, 0.036] |
| Inferior Occipital Gyrus | 1.02 [0.96, 1.03] | 1.07 [1.04, 1.10] | 0.020 [-0.013, 0.059] | -0.006 [-0.031, 0.023] |
| Cuneus | 0.99 [0.93, 1.01] | 1.00 [0.96, 1.04] | -0.018 [-0.046, 0.014] | 0.002 [-0.015, 0.048] |
| Lingual Gyrus | 0.97 [0.95, 1.02] | 0.97 [0.93, 1.03] | -0.000 [-0.016, 0.033] | -0.011 [-0.029, 0.030] |
| Cingulate Gyrus | 0.96 [0.92, 1.01] | 0.94 [0.92, 0.99] | 0.005 [-0.003, 0.014] | 0.005 [-0.008, 0.013] |
| Insular Cortex | 0.92 [0.87, 0.95] | 0.92 [0.88, 0.97] | -0.000 [-0.018, 0.021] | -0.010 [-0.028, 0.008] |
| Amygdala | 0.99 [0.97, 1.05] | 1.02 [0.99, 1.11] | 0.005 [-0.015, 0.037] | 0.018 [-0.005, 0.047] |
| Hippocampus | 1.56 [1.15, 2.08] | 1.44 [1.23, 1.66] | 0.001 [-0.140, 0.256] | -0.009 [-0.076, 0.045] |
| Caudate Nucleus | 0.91 [0.81, 0.95] | 0.95 [0.87, 1.02] | -0.009 [-0.075, 0.020] | 0.018 [-0.030, 0.063] |
| Putamen | 1.13 [1.04, 1.19] | 1.30 [1.21, 1.36] | -0.002 [-0.035, 0.023] | 0.008 [-0.001, 0.043] |
| Globus Pallidus | 1.27 [1.20, 1.33] | 1.57 [1.40, 1.84] | -0.009 [-0.048, 0.026] | 0.034 [0.010, 0.103] |
| Thalamus | 1.50 [1.40, 1.73] | 1.69 [1.50, 1.85] | 0.015 [-0.070, 0.061] | 0.040 [-0.002, 0.091] |
| Basal Forebrain | 1.06 [1.01, 1.11] | 1.16 [1.07, 1.26] | -0.005 [-0.018, 0.062] | -0.008 [-0.039, 0.027] |
| Nucleus Accumbens | 1.01 [0.97, 1.05] | 1.13 [1.04, 1.19] | -0.007 [-0.030, 0.033] | -0.024 [-0.060, 0.017] |
| Midbrain | 1.24 [1.21, 1.33] | 1.51 [1.37, 1.68] | 0.010 [-0.032, 0.021] | 0.004 [-0.047, 0.053] |
| Cerebellum Gray Matter | 1.10 [1.08, 1.17] | 1.11 [1.07, 1.18] | 0.015 [-0.048, 0.045] | 0.014 [-0.016, 0.030] |
| Pons | 1.29 [1.17, 1.43] | 1.24 [1.12, 1.31] | -0.021 [-0.044, 0.078] | 0.011 [-0.023, 0.052] |
| Medulla | 1.13 [1.00, 1.26] | 1.07 [0.90, 1.15] | -0.031 [-0.140, 0.111] | 0.001 [-0.055, 0.068] |
| Anterior Deep White Matter | 1.05 [0.95, 1.14] | 0.96 [0.89, 1.04] | -0.011 [-0.047, 0.019] | 0.008 [-0.045, 0.059] |
| Posterior Deep White Matter | 0.98 [0.88, 1.10] | 0.85 [0.79, 0.95] | -0.008 [-0.078, 0.045] | -0.007 [-0.059, 0.050] |
| Genu of the Corpus Callosum | 0.94 [0.86, 1.02] | 0.83 [0.75, 0.90] | -0.016 [-0.045, 0.038] | 0.002 [-0.052, 0.036] |
| Body of the Corpus Callosum | 0.91 [0.80, 1.07] | 0.84 [0.74, 0.89] | -0.023 [-0.098, 0.102] | -0.040 [-0.080, 0.023] |
| Splenium of the Corpus Callosum | 0.97 [0.90, 1.08] | 0.85 [0.77, 0.91] | -0.024 [-0.060, 0.025] | -0.027 [-0.084, 0.035] |
| Periventricular White Matter (lateral) | 0.86 [0.67, 0.97] | 0.65 [0.58, 0.77] | -0.004 [-0.109, 0.040] | -0.005 [-0.089, 0.065] |
| Anterior Limb of the Internal Capsule | 1.18 [1.05, 1.21] | 1.25 [1.15, 1.33] | -0.014 [-0.105, 0.023] | 0.046 [-0.021, 0.088] |
| Posterior Limb of the Internal Capsule | 1.23 [1.14, 1.26] | 1.26 [1.16, 1.39] | -0.026 [-0.070, 0.005] | 0.026 [-0.023, 0.132] |
| Inferior Deep White Matter | 1.13 [1.09, 1.18] | 1.03 [0.98, 1.08] | -0.037 [-0.081, -0.002] | 0.012 [-0.051, 0.047] |
| Cingulum (Cingulate Gyrus segment) | 1.07 [1.04, 1.11] | 1.01 [0.97, 1.07] | -0.023 [-0.063, 0.028] | 0.011 [-0.028, 0.034] |
| Cingulum (Hippocampal Segment) | 1.05 [1.01, 1.17] | 1.03 [0.97, 1.10] | -0.002 [-0.045, 0.058] | 0.012 [-0.030, 0.038] |
| Fornix / Stria Terminalis | 1.92 [1.46, 2.27] | 1.63 [1.39, 1.85] | 0.025 [-0.138, 0.133] | 0.023 [-0.069, 0.083] |
| Fornix | 1.20 [0.91, 1.53] | 0.82 [0.67, 1.05] | 0.025 [-0.133, 0.157] | -0.008 [-0.081, 0.046] |
| Peripheral Parietal White Matter | 1.03 [1.01, 1.08] | 1.04 [0.99, 1.08] | -0.011 [-0.049, 0.020] | 0.011 [-0.012, 0.044] |
| Periventricular White Matter (anterior) | 0.81 [0.72, 0.96] | 0.70 [0.64, 0.80] | -0.027 [-0.063, 0.081] | -0.022 [-0.054, 0.020] |
| Periventricular White Matter (posterior) | 1.73 [1.30, 2.08] | 1.24 [1.00, 1.70] | -0.007 [-0.298, 0.048] | 0.036 [-0.050, 0.087] |
| Peripheral Frontal White Matter | 1.06 [1.02, 1.09] | 1.08 [1.02, 1.15] | -0.002 [-0.015, 0.010] | 0.016 [-0.009, 0.038] |
| Peripheral Temporal White Matter | 1.06 [1.05, 1.08] | 1.05 [1.01, 1.09] | -0.014 [-0.036, 0.012] | -0.003 [-0.015, 0.033] |
| Peripheral Occipital White Matter | 1.09 [1.05, 1.12] | 1.08 [1.03, 1.12] | -0.012 [-0.028, 0.012] | -0.001 [-0.011, 0.019] |
| Peripheral Cingulate White Matter | 1.10 [1.07, 1.14] | 1.03 [1.00, 1.10] | -0.012 [-0.038, -0.002] | 0.013 [-0.018, 0.044] |
| Cerebellum White Matter | 1.39 [1.35, 1.49] | 1.34 [1.28, 1.45] | -0.023 [-0.100, 0.019] | 0.029 [-0.017, 0.061] |
| Caudate tail | 2.14 [1.61, 3.02] | 1.88 [1.46, 2.20] | -0.013 [-0.336, 0.160] | 0.014 [-0.063, 0.110] |

Values are expressed as median [interquartile range].
Abbreviations: HC, healthy controls; PSP, progressive supranuclear palsy; SUVR, Standardized uptake value ratio; ΔSUVR, Annualized change in standardized uptake value ratio

**Table S3.** Summary of the fixed effects in the linear mixed-effects models across key regions

| **Regions** | **Fixed effects** | **β [95% Confidence Interval]** | **p-value** |
| --- | --- | --- | --- |
| Globus Pallidus | PSP rating scale | 0.0043 [0.0008, 0.0078] | **0.018** |
|  | Timing | 0.0087 [-0.0289, 0.0463] | 0.641 |
| Midbrain | PSP rating scale | 0.0026 [-0.0008, 0.0059] | 0.134 |
|  | Timing | -0.0105 [-0.0471, 0.0262] | 0.564 |
| Superior Parietal Gyrus | PSP rating scale | -0.0009 [-0.0008, 0.0025] | 0.286 |
|  | Timing | 0.0344 [0.0157, 0.0531] | **<0.001** |
| Cerebellar White Matter | PSP rating scale | 0.0028 [-0.0002, 0.0057] | 0.052 |
|  | Timing | 0.0069 [-0.0240, 0.0377] | 0.756 |
| Pons | PSP rating scale | 0.0030 [0.0003, 0.0058] | **0.033** |
|  | Timing | -0.0046 [-0.0337, 0.0246] | 0.751 |

Bold values indicate statistical significance (p < 0.05).

Abbreviations: PSP, progressive supranuclear palsy.

**Table S4.** Preliminary validation comparing longitudinal tau accumulation with and without partial volume correction

| **Regions** | **ΔSUVR  without PVC** | **ΔSUVR  with PVC** |
| --- | --- | --- |
| Globus Pallidus | 0.034 [0.010, 0.103]** | 0.073 [0.002, 0.225]** |
| Midbrain | 0.004 [-0.047, 0.053] | 0.017 [-0.028, 0.085] |
| Superior Parietal Gyrus | 0.030 [0.014, 0.047]*** | 0.033 [-0.015, 0.129]* |
| Cerebellar White Matter | 0.029 [-0.017, 0.061]* | 0.045 [-0.024, 0.123]* |

Values represent the annualized change in SUVR (median [interquartile range]).

Significant longitudinal increase compared with baseline (Wilcoxon signed-rank test): **p* < 0.05, ***p* < 0.01, ****p* < 0.001

Abbreviations: ΔSUVR, Annualized change in standardized uptake value ratio; PVC, Partial volume correction.

**Table S5.** Volumetric and longitudinal tracer uptake characteristics across three reference regions in healthy controls and progressive supranuclear palsy

|  | **HC** | **PSP** |  |  | ***p*-value** |  |
| --- | --- | --- | --- | --- | --- | --- |
|  |  | **All** | **Lower severity** | **Higher severity** | **PSP vs. HC** | **LS vs. HS** |
| **GM-ref** |  |  |  |  |  |  |
| Volume (cm^3^) | 148.1 [108.8, 201.9] * | 114.8 [101.0, 136.6] * | 111.8 [98.1, 156.9] | 122.3 [100.2, 126.8] * | 0.143 | 0.682 |
| Baseline SUV (g/ml) | 0.324 [0.297, 0.369] | 0.336 [0.262, 0.405] | 0.375 [0.257, 0.402] | 0.308 [0.265, 0.420] | 0.711 | 1.000 |
| **Δ**SUV (g/ml) | 0.005 [-0.053, 0.023] | -0.003 [-0.028, 0.043] | 0.010 [-0.043, 0.051] | -0.004 [-0.013, 0.042] | 0.537 | 0.838 |
| *p*-value (**Δ**SUV vs. 0) | 0.922 | 0.648 | 0.839 | 0.946 | — | — |
| **WM-ref** |  |  |  |  |  |  |
| Volume (cm^3^) | 143.2 [116.4, 176.4] * | 122.0 [103.0, 156.4] * | 139.7 [98.9, 172.0] * | 119.1 [102.2, 147.0] * | 0.223 | 0.538 |
| Baseline SUV (g/ml) | 0.331 [0.321, 0.361] | 0.341 [0.270, 0.398] | 0.366 [0.263, 0.399] | 0.319 [0.288, 0.385] | 0.818 | 0.838 |
| **Δ**SUV (g/ml) | -0.003 [-0.059, 0.028] | 0.005 [-0.020, 0.033] | 0.012 [-0.022, 0.044] | 0.003 [-0.015, 0.027] | 0.331 | 0.838 |
| *p*-value (**Δ**SUV vs. 0) | 0.770 | 0.313 | 0.376 | 0.636 | — | — |
| **CB-ref** |  |  |  |  |  |  |
| Volume (cm^3^) | 89.8 [86.0, 105.6] | 98.7 [89.6, 104.3] | 99.6 [90.2, 106.4] | 93.6 [88.7, 102.3] | 0.448 | 0.383 |
| Baseline SUV (g/ml) | 0.365 [0.331, 0.399] | 0.361 [0.286, 0.450] | 0.390 [0.284, 0.457] | 0.361 [0.289, 0.455] | 0.818 | 0.918 |
| **Δ**SUV (g/ml) | -0.014 [-0.062, 0.043] | 0.005 [-0.031, 0.063] | 0.019 [-0.028, 0.068] | -0.001 [-0.037, 0.048] | 0.298 | 0.573 |
| *p*-value (**Δ**SUV vs. 0) | 0.557 | 0.406 | 0.305 | 1.000 | — | — |

Values are expressed as median [interquartile range]. Lower and higher severity groups were defined by the median PSP Rating Scale score.

* *p* < 0.05 compared with CB-ref (Wilcoxon signed-rank test)

Abbreviations: CB-ref, Cerebellar reference; GM-ref, Histogram-based gray matter reference; HC, Healthy control; HS, Higher severity; LS, Lower severity; PSP, progressive supranuclear palsy; WM-ref, Histogram-based white matter reference; ΔSUV, Annualized change in standardized uptake value;

**Table S6.** Comparison of baseline and longitudinal SUVRs among three reference regions

|  | **GM-ref** | **WM-ref** | **CB-ref** |
| --- | --- | --- | --- |
| **Globus Pallidus** |  |  |  |
| Baseline SUVR |  |  |  |
| PSP | 1.57 [1.40, 1.84] | 1.54 [1.44, 1.73] | 1.43 [1.27, 1.64] |
| HC | 1.27 [1.20, 1.33] | 1.18 [1.10, 1.24] | 1.13 [1.09, 1.19] |
| *p*-value (PSP vs. HC) | **<0.001** | **<0.001** | **0.001** |
| AUC | 0.888 | 0.946 | 0.862 |
| Annual changes in SUVR |  |  |  |
| PSP | 0.034 [0.010, 0.103] | 0.042 [0.004, 0.089] | 0.040 [-0.027, 0.079] |
| HC | -0.009 [-0.048, 0.026] | 0.004 [-0.023, 0.046] | -0.029 [-0.069, 0.046] |
| *p*-value (PSP vs. 0) | **0.003** | **0.002** | 0.082 |
| *p*-value (PSP vs. HC) | **0.016** | 0.108 | 0.153 |
| AUC | 0.765 | 0.677 | 0.658 |
| Correlation between **Δ**SUVR and **Δ**PSP Rating Scale scores in PSP-RS | | | |
| Spearman’s ρ | 0.688 | 0.344 | 0.511 |
| *p*-value | **0.002** | 0.162 | **0.030** |
| **Midbrain** |  |  |  |
| Baseline SUVR |  |  |  |
| PSP | 1.51 [1.37, 1.68] | 1.51 [1.38, 1.62] | 1.34 [1.21, 1.48] |
| HC | 1.24 [1.21, 1.33] | 1.17 [1.10, 1.30] | 1.12 [1.11, 1.14] |
| *p*-value (PSP vs. HC) | **0.001** | **<0.001** | **<0.001** |
| AUC | 0.850 | 0.908 | 0.881 |
| Annual changes in SUVR |  |  |  |
| PSP | 0.004 [-0.047, 0.053] | 0.019 [-0.014, 0.055] | 0.008 [-0.061, 0.039] |
| HC | 0.010 [-0.032, 0.021] | 0.026 [-0.010, 0.071] | 0.006 [-0.034, 0.066] |
| *p*-value (PSP vs. 0) | 0.420 | 0.215 | 0.853 |
| *p*-value (PSP vs. HC) | 0.902 | 0.791 | 0.560 |
| AUC | 0.485 | 0.531 | 0.565 |
| **Superior Parietal Gyrus** |  |  |  |
| Baseline SUVR |  |  |  |
| PSP | 0.92 [0.85, 0.98] | 0.90 [0.85, 0.98] | 0.80 [0.75, 0.88] |
| HC | 0.83 [0.81, 0.92] | 0.79 [0.73, 0.87] | 0.76 [0.67, 0.84] |
| *p*-value (PSP vs. HC) | **0.036** | **0.010** | 0.251 |
| AUC | 0.731 | 0.781 | 0.627 |
| Annual changes in SUVR |  |  |  |
| PSP | 0.030 [0.014, 0.047] | 0.030 [-0.006, 0.070] | 0.022 [-0.012, 0.052] |
| HC | -0.007 [-0.019, 0.010] | 0.018 [-0.027, 0.035] | 0.007 [-0.041, 0.024] |
| *p*-value (PSP vs. 0) | **<0.001** | **0.007** | **0.020** |
| *p*-value (PSP vs. HC) | **0.002** | 0.266 | 0.153 |
| AUC | 0.835 | 0.623 | 0.658 |
| **Cerebellum white matter** |  |  |  |
| Baseline SUVR |  |  |  |
| PSP | 1.34 [1.28, 1.45] | 1.35 [1.25, 1.43] | 1.21 [1.13, 1.32] |
| HC | 1.39 [1.35, 1.49] | 1.33 [1.22, 1.45] | 1.26 [1.21, 1.34] |
| *p*-value (PSP vs. HC) | 0.163 | 0.986 | 0.266 |
| AUC | 0.653 | 0.504 | 0.623 |
| Annual changes in SUVR |  |  |  |
| PSP | 0.029 [-0.017, 0.061] | 0.019 [-0.003, 0.053] | -0.002 [-0.021, 0.056] |
| HC | -0.023 [-0.100, 0.019] | 0.004 [-0.059, 0.045] | -0.049 [-0.079, 0.007] |
| *p*-value (PSP vs. 0) | **0.016** | **0.011** | 0.465 |
| *p*-value (PSP vs. HC) | **0.021** | 0.101 | 0.054 |
| AUC | 0.754 | 0.681 | 0.712 |

Values are expressed as median [interquartile range]. Bold values indicate statistical significance (p < 0.05).

Abbreviations: AUC, Area under the curve; CB-ref, Cerebellar reference; GM-ref, Histogram-based gray matter reference; HC, Healthy control; PSP, progressive supranuclear palsy; SUVR, Standardized uptake value ratio; WM-ref, Histogram-based white matter reference; ΔSUVR, Annualized change in standardized uptake value ratio.

**Figure S1.** Reference region definition strategies for longitudinal quantification

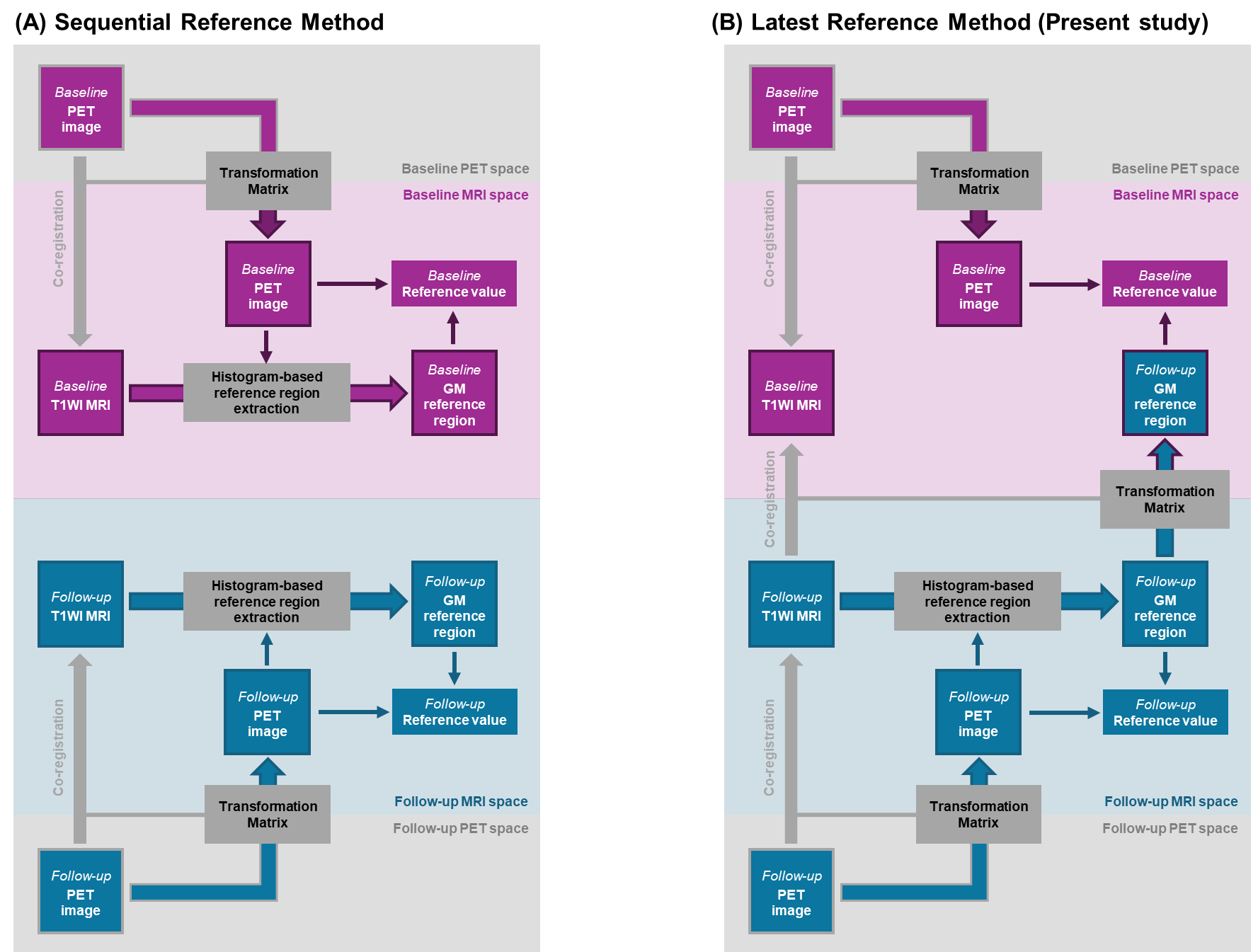

(A) Sequential reference method. Reference regions are independently defined at each time point based on signal intensity histograms. Although this approach reflects time-specific tracer distributions, it may introduce artificial longitudinal variability owing to progressive pathology or tissue atrophy affecting the reference tissue itself. (B) Latest reference method (present study). A histogram-based reference region is defined on the follow-up image and spatially back-transformed to the baseline image, thereby ensuring anatomical and voxelwise consistency across time points. The follow-up scan was chosen as the standard space as tissue with persistently low tracer retention at more advanced disease stages is expected to provide the most stable and conservative reference for longitudinal quantification. Box fill colors denote the time point of data acquisition (purple: baseline; blue: follow-up). Box border colors indicate the space of the image co-registered (purple: baseline space; blue: follow-up space).

**Figure S2.** Individual longitudinal changes in PSP Rating Scale scores

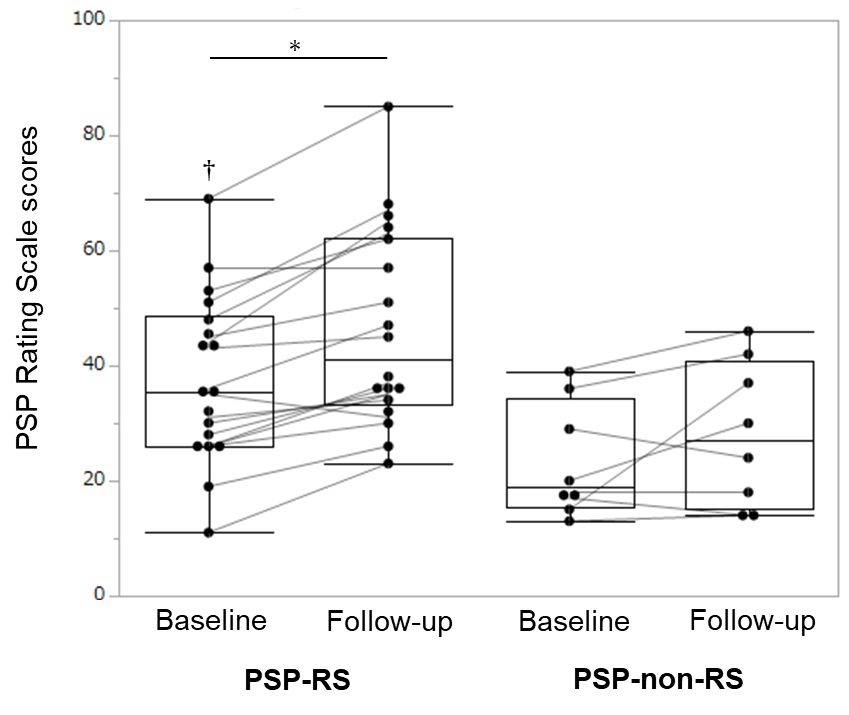

PSP Rating Scale scores at baseline and follow-up for patients with Richardson‘s syndrome (PSP-RS) and non-RS variants (PSP-non-RS). Individual trajectories are connected by lines. The asterisk denotes a significant longitudinal increase compared with baseline (*p* < 0.05, Wilcoxon signed-rank test). Daggers indicate significant differences between PSP-RS and non-RS at baseline (*p* < 0.05, Mann–Whitney U test).

Abbreviations: PSP-RS, progressive supranuclear palsy Richardson’s syndrome; PSP-non-RS, progressive supranuclear palsy without Richardson’s syndrome.

**Figure S3.** Diagnostic performance of baseline SUVR and ΔSUVR for differentiating patients with PSP from healthy controls

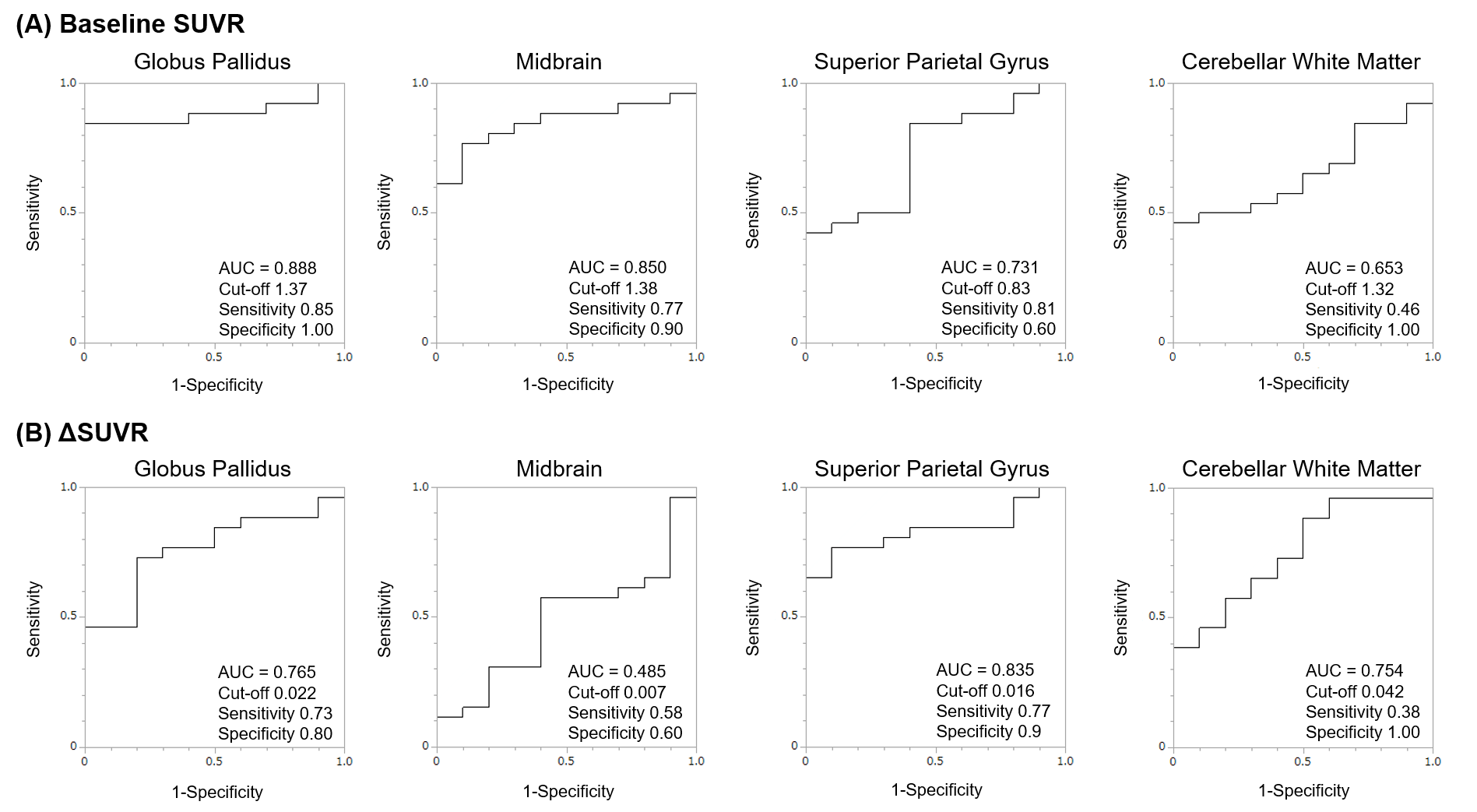

Receiver operating characteristic (ROC) curves illustrate the diagnostic accuracy of (A) baseline SUVR and (B) annual longitudinal SUVR changes (ΔSUVR) in distinguishing patients with PSP from healthy controls. Data are shown for the following four representative regions: the globus pallidus, midbrain, superior parietal gyrus, and cerebellar white matter. For each plot, the area under the curve (AUC), optimal cut-off value, sensitivity, and specificity are provided.

Abbreviations: AUC, area under the curve; HC, healthy controls; PSP, progressive supranuclear palsy; ROC, receiver operating characteristic; SUVR, standardized uptake value ratio; ΔSUVR, annualized change in standardized uptake value ratio.

**Figure S4.** Spatial distribution of histogram-based reference regions in patients with PSP and healthy controls

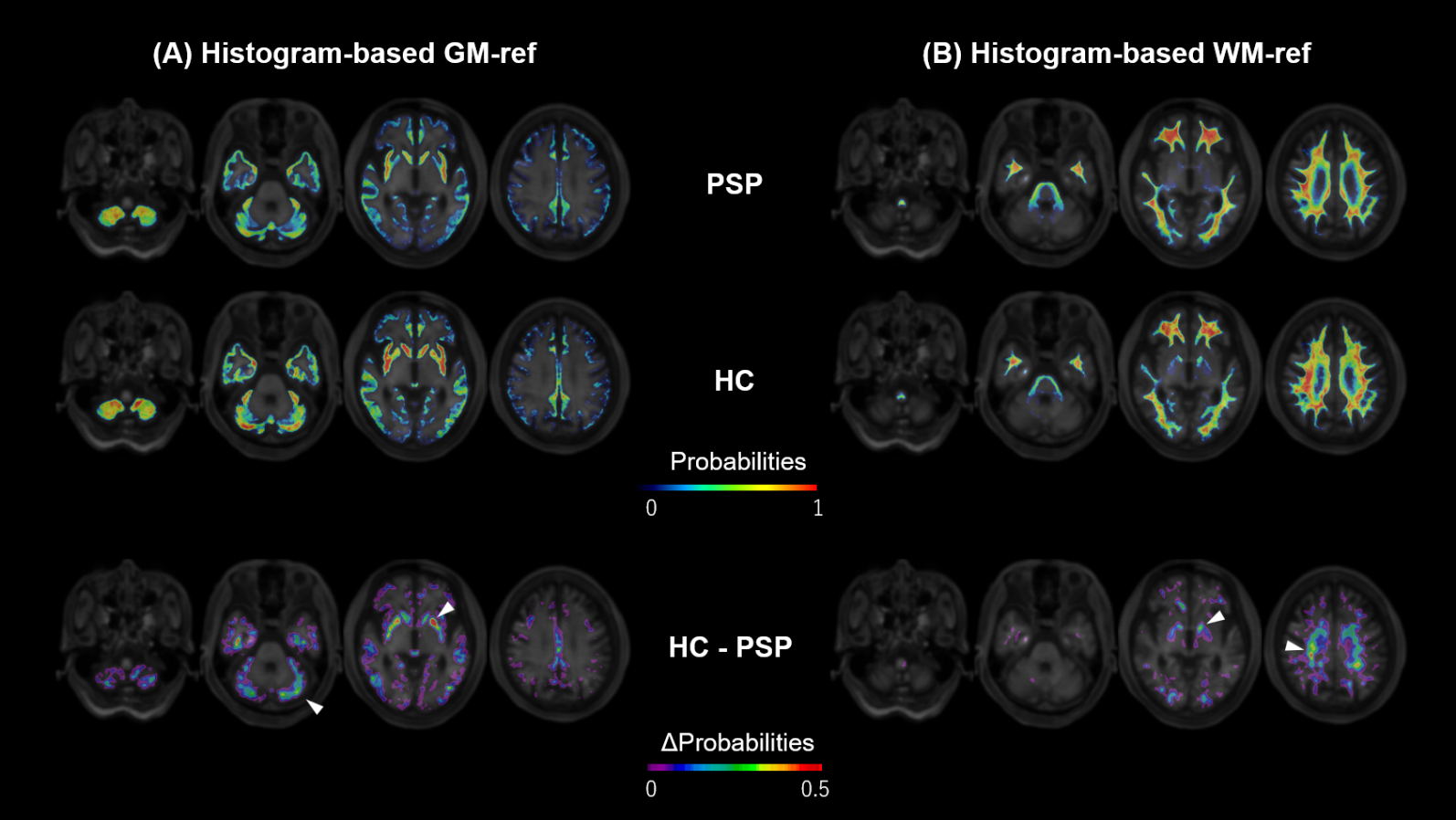

Group-averaged probability maps for **(A)** histogram-based gray matter (GM-ref) and **(B)** white matter (WM-ref) reference regions. The WM-ref was defined by substituting white matter for gray matter using the same algorithm described in Figure S1.The top and middle rows display the spatial distribution of these regions in patients with progressive supranuclear palsy (PSP) and healthy controls (HC), respectively. The bottom row illustrates the difference in probabilities between the two groups (HC - PSP), smoothed with a 3-mm filter for visualization purposes. The upper color bar indicates the probability values ranging from 0 to 1, whereas the lower color bar represents the difference in probabilities (ΔProbabilities) ranging from 0 to 0.5. White arrowheads highlight specific regions exhibiting relatively large intergroup differences, such as the cerebellar gray matter, basal ganglia, and the subcortical white matter, likely reflecting the exclusion of regions with disease-related tau accumulation by the histogram-based approach.

Abbreviations: GM-ref, gray matter reference; HC, healthy controls; PSP, progressive supranuclear palsy; WM-ref, white matter reference.

**Figure S5.** Longitudinal changes in SUVR and ΔSUVR using histogram-based white matter reference region

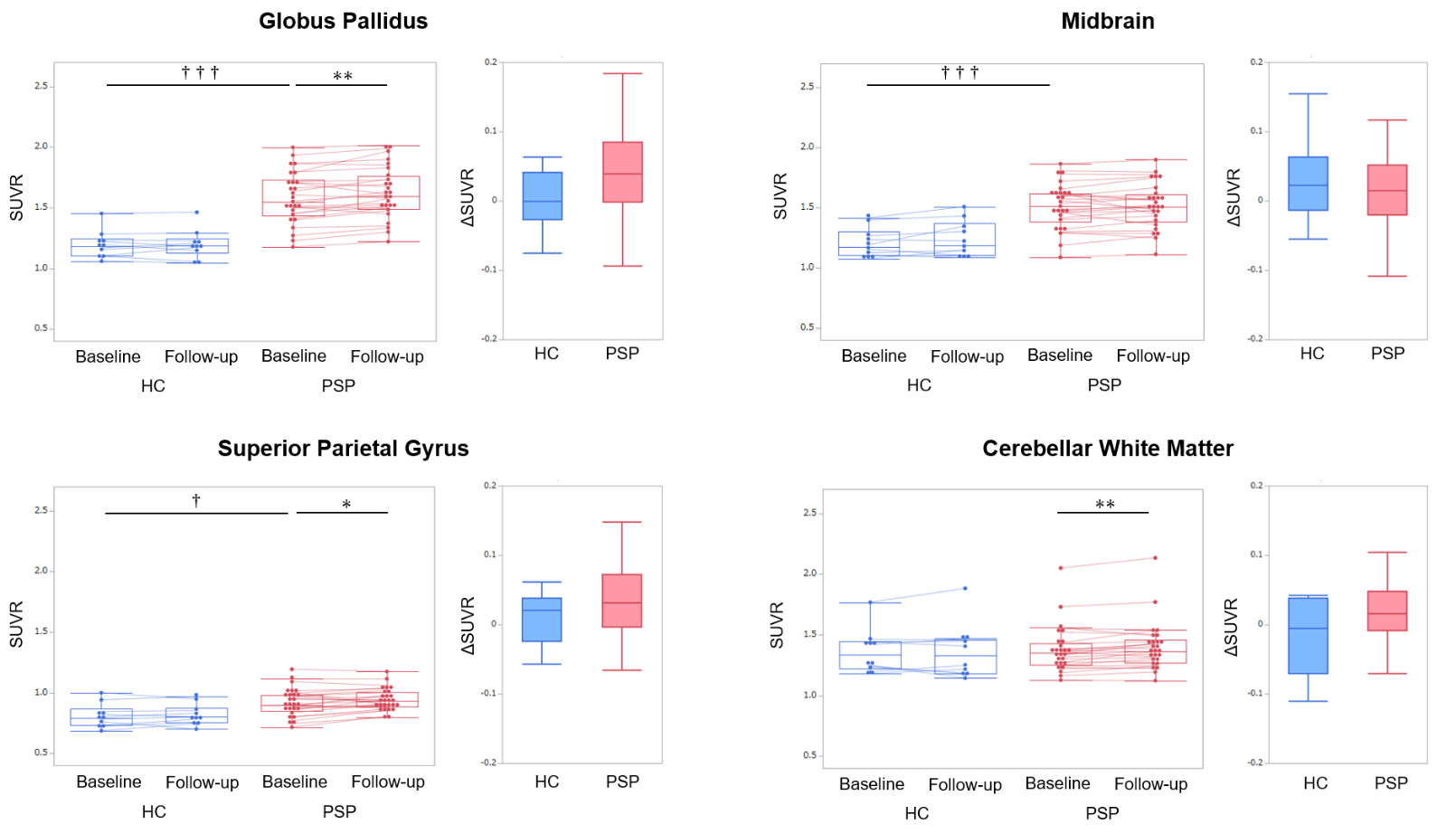

Pairs of graphs are displayed for four key regions. For each region, comparison of standardized uptake value ratios between baseline and follow-up for healthy controls (HCs, blue) and patients with progressive supranuclear palsy (PSP, red) in the left graphs, with individual trajectories are connected by lines. Direct group comparisons of annualized ΔSUVR between the HCs and PSP groups are shown in the right graphs, with no significant group differences observed. Asterisks denote significant longitudinal changes within each cohort (* p < 0.05, ** p < 0.01, *** p < 0.001). Daggers indicate significant differences between HCs and PSP in either baseline SUVR or ΔSUVR († p < 0.05, †† p < 0.01, ††† p < 0.001).

Abbreviations: SUVR, standardized uptake value ratio; ΔSUVR, annualized change in standardized uptake value ratio; HCs, healthy controls; PSP, progressive supranuclear palsy.

**Figure S6.** Association between longitudinal tau accumulation and clinical progression in PSP-RS using histogram-based white matter reference region

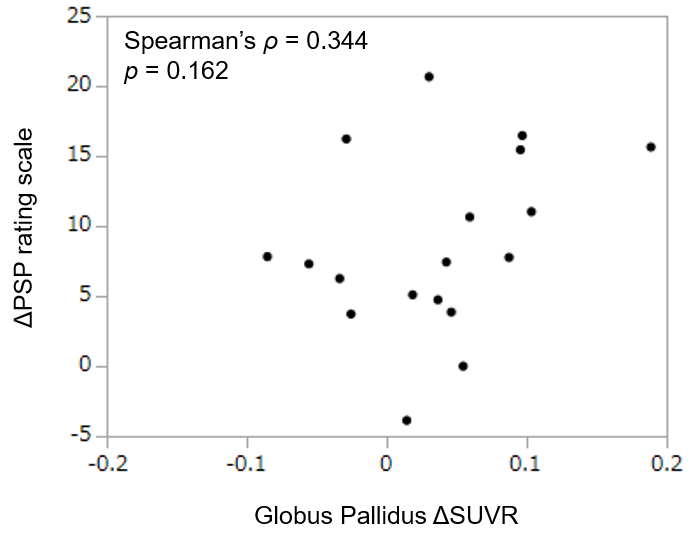

Scatter plot showing the association between ΔSUVR in the GP and ΔPSP Rating Scale in PSP-RS, calculated using the histogram-based white matter reference region.

Abbreviations: GP, globus pallidus; PSP-RS, progressive supranuclear palsy Richardson’s syndrome; SUVR, standardized uptake value ratio;ΔSUVR, annualized change in standardized uptake value ratio.

**Figure S7** Effect of reference region selection on ΔSUVR variability across tissue types

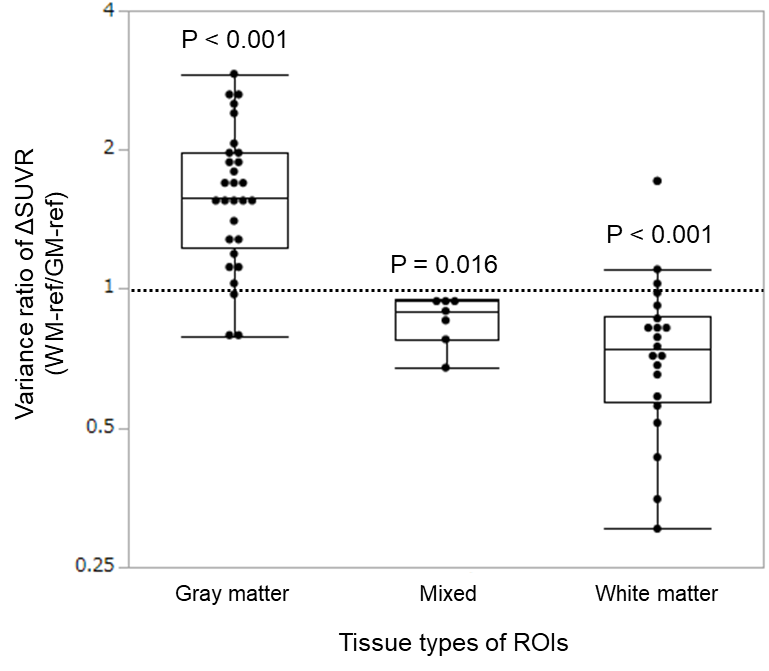

Variance ratio of ΔSUVR (variance of ΔSUVR with WM-ref divided by that with GM-ref) across ROIs categorized by tissue type. The variance of ΔSUVR was calculated in healthy controls (HCs, n = 10) using white and gray matter reference regions, and the ratio of these variances was determined for each ROI. ROIs were classified into gray matter, white matter, and mixed tissue regions. The mixed category included regions with gray and white matter components (globus pallidus, thalamus, basal forebrain, hippocampus, midbrain, pons, and medulla). The y-axis denotes the base-2 logarithm of this variance ratio. P values were derived using the Wilcoxon signed-rank test to determine whether the variance ratio differed from 1. These results demonstrate that longitudinal variance significantly increases when the tissue type of the target ROI and reference region are mismatched.

Abbreviations: ΔSUVR, annualized change in standardized uptake value ratio; WM-ref, histogram-based white matter reference; GM-ref, histogram-based gray matter reference; ROI, region of interest; HC, healthy control.

**Figure S8.** Effect of disease severity on spatial distribution of histogram-based reference regions in PSP.

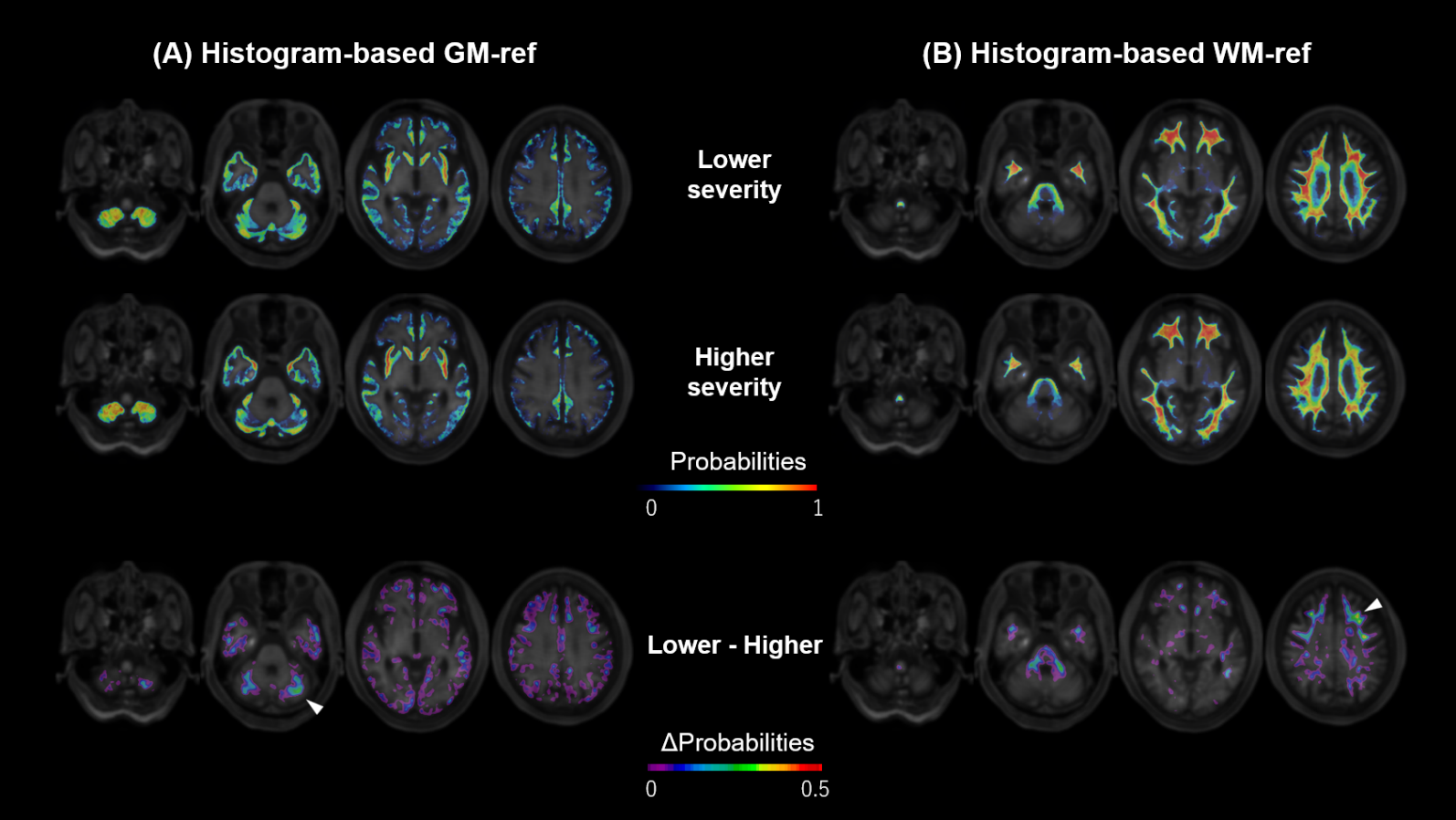

Group-averaged probability maps for **(A)** histogram-based gray matter (GM-ref) and **(B)** white matter (WM-ref) reference regions in progressive supranuclear palsy (PSP) stratified by disease severity. The PSP cohort was divided into two groups (n = 13, respectively) based on the median PSP Rating Scale score at follow-up. The top and middle rows depict the spatial distribution of these regions in the lower and higher severity PSP groups, respectively. The bottom row illustrates the difference in probabilities between the two groups (Lower severity − Higher severity), smoothed with a 3-mm filter for visualization purposes. The upper color bar indicates the probability values ranging from 0 to 1, whereas the lower color bar represents the difference in probabilities (ΔProbabilities) ranging from 0 to 0.5. White arrowheads highlight specific regions exhibiting relatively large differences between the severity groups, such as the cerebellar gray matter and frontal white matter, possibly reflecting the further exclusion of regions with advancing disease-related tau accumulation in the higher severity group.

Abbreviations: GM-ref, gray matter reference; PSP, progressive supranuclear palsy; WM-ref, white matter reference.

**Supplementary Method**

*Radioligand synthesis*

Radiosynthesis of florzolotau and [11C]PiB was performed as previously described.^1-3^ Florzolotau was synthesized through the reaction of its tosylate precursor with [18F]fluoride in the presence of dimethyl sulfoxide, K_2_CO_3_, and K222. The resulting product was purified via analytic high-performance liquid chromatography (Waters Atlantis prep T3 column, 4.6 × 150 mm; CH_3_CN/50 mM AcONH_4_ = 4/6, 1 mL/min). The final formulation achieved a radiochemical purity of at least 95%. The specific activities at the time of injection were 140.4–396.3 GBq/µmol (baseline) and 51.7–504.2 GBq/µmol (follow-up) for florzolotau and 52.2–340.8 GBq/µmol for [11C]PiB.

*Partial Least Square (PLS) analysis*

PLS analyses were conducted using JMP Pro 18 (SAS Institute Inc., Cary, NC, USA). PLS model estimation was performed using the nonlinear iterative partial least squares algorithm.^4^ The optimal number of latent components was determined to minimize the predicted residual sum of squares using leave-one-out cross-validation, defined as follows:

$$PRESS= \sum_{i=1}^{n} {(y_{i}-\hat{y}_{i,-i})}^{2}$$

where *y_i_* denotes the observed value of the response variable for the *i*-th subject, and $\hat{y}_{i,-i}$ is the predicted value obtained from a model constructed without the *i*-th subject’s data.^5^ Model performance was assessed based on the cumulative proportion of variation explained in the response variable (*R^2^Y*) and the cross-validated predictive ability (*Q²*),^6^ defined as follows:

$$R^{2}Y= 1-\frac{RSS}{TSS}$$

$$Q^{2}=1-\frac{PRESS}{TSS}$$

where $RSS=\sum_{i=1}^{n} {(y_{i}-\hat{y}_{i})}^{2}$ is the residual sum of squares; $TSS=\sum_{i=1}^{n} {(y_{i}-\bar{y})}^{2}$, the total sum of squares of the response variable; $\hat{y}_{i}$, the predicted value of the response variable for the *i*-th subject; and $\bar{y}$, the mean value of the response variable.^7^ A *Q²* value greater than zero was considered to indicate that the model had meaningful predictive ability in cross-validation, demonstrating superior performance to prediction based on the mean response. To assess the contribution of individual ROIs to the PLS model, Variable Importance in Projection (VIP) scores and regression coefficients were calculated. The VIP score for the *j*-th predictor was defined as follows:

$${VIP}_{j}=\sqrt{\frac{p\sum_{a=1}^{A} w_{a,j}^{2}{R^{2}Y}_{a}}{\sum_{a=1}^{A} {R^{2}Y}_{a}}}$$

where *p* denotes the number of predictor variables; *A*, the total number of latent components; *w_a,j_*, the weight of the *j*-th predictor for the *a*-th latent component; and *R^2^Y_a_*, the variation of *Y* explained by the *a*-th latent component. Primary contributors were defined as ROIs with VIP > 1.5 and positive regression coefficients. Although VIP > 1 is generally regarded as reflecting contribution to PLS models, a more stringent threshold was applied to enhance regional specificity and interpretability.^8^

**Supplementary Discussion**

To comprehensively assess the optimal reference region for longitudinal assessments, three approaches were compared: the primary histogram-based gray matter reference region (GM-ref), a histogram based white matter reference region (WM-ref), and a conventional fixed cerebellar gray matter reference region (CB-ref). The WM-ref was defined by substituting white matter for gray matter using the same histogram-based algorithm described for the GM-ref (Figure S1), whereas the CB-ref was selected from our predefined set of regions of interest (ROIs).

The volumes of the histogram-based references were significantly larger than that of the fixed CB-ref and did not significantly differ between PSP and HCs (Table S5), which generally helps reduce the impact of localized noise and improve measurement robustness. Regarding longitudinal stability, none of the three reference regions showed significant longitudinal changes in SUV in either group (Table S5), suggesting no apparent systematic drift over the 1-year follow-up period. However, variability in raw SUV measurements may have limited the sensitivity for detecting subtle systematic alterations.^9^

To explore the influence of disease severity on the histogram-based reference region selection, the PSP cohort was divided into two groups based on the median PSP Rating Scale scores. Although no significant differences were observed in the total volumes of the GM-ref or WM-ref between the two groups, their spatial distributions exhibited regional differences. Specifically, cerebellar gray matter and frontal white matter were less frequently selected in the higher severity group, possibly reflecting increased tau accumulation in these regions with disease progression (Fig. S8).

Subsequently, the effect of reference region selection on the longitudinal measurement of SUVR in target regions was assessed (Table S6). When the CB-ref was used, the longitudinal increase in target SUVR observed with the GM-ref was markedly attenuated, specifically in group comparisons and several key ROIs. Such a reduced sensitivity may reflect subtle tau accumulation within the cerebellar reference region during disease progression, thus reducing the target-to-reference contrast. Likewise, with the WM-ref, although a longitudinal increase in SUVR was observed within the PSP group, significant group differences were not observed between PSP and HCs. This lack of disease specificity indicates that longitudinal changes inherent to white matter, including previously reported age-related changes,^10^ may mask true disease-related changes in SUVR.

Furthermore, the WM-ref failed to demonstrate the correlation between longitudinal tau accumulation in the globus pallidus and clinical progression in PSP Rating Scale scores (Table S6). To explore the basis for this reduced sensitivity, analysis of ΔSUVR variance in HCs revealed that measurement stability depended on tissue matching. The variance was greater when tissue types were mismatched between target ROIs and reference regions (e.g., a gray matter target paired with the WM-ref) than when they were matched (Fig. S7). Such an increased variance with tissue mismatch likely reflects differences in tracer retention and nonspecific binding between gray and white matter.^11^ Supporting this interpretation, the CB-ref—a conventional gray matter reference—also exhibited clinical correlation, consistent with the GM-ref.

Collectively, these findings support the utility of the histogram-based gray matter reference adopted in this study for longitudinal tau PET assessments in PSP and indicate that tissue matching between reference and target regions is crucial for measurement sensitivity optimization, particularly when evaluating the predominantly gray matter regions identified in our analyses. Conversely, white matter reference regions may remain appropriate for the assessment of white matter target regions, although this requires further validation. These observations also raise the possibility that tissue-specific or mixed-tissue reference approaches provide a more flexible normalization strategy across different target regions, warranting investigation in larger multicenter cohorts.
